# A method for robust estimation of seasonal onset and intensity in respiratory surveillance data: evaluated using data from 21 European countries

**DOI:** 10.1101/2025.11.18.25340240

**Authors:** Sofia Myrup Otero, Hanne-Dorthe Emborg, Kasper Schou Telkamp, Ida Rask Moustsen-Helms, Bolette Søborg, Lasse Engbo Christiansen

## Abstract

**Background:** Seasonal respiratory pathogens cause hospital admissions and strain health services. Commonly used methods, Moving Epidemic Method (mem), WHO Average Curve Method (WHO-ACM) and Mean Standard Deviation method (MSD), support post-season intensity assessment. We developed aedseo for early seasonal onset detection and within-season intensity assessment.

**Methods:** aedseo fits a rolling quasi-Poisson generalised linear model to weekly counts; onset is declared when growth is significant and the five-week mean exceeds a disease-specific threshold (*T*_*disease*_) estimated from recent seasons. Intensity breakpoints (between intensity levels; very low to very high) are disease-specific; *T*_*disease*_ defines ‘very low’; ‘high’ is the 97.5th percentile of a log-normal fitted to three highest weekly counts per season; ‘low’ and ‘medium’ are equally log-spaced in-between. aedseo was developed on Danish respiratory surveillance data and validated with 63 data sources (influenza, RSV, ARI, ILI) from 21 European countries (seasons 2014/15–2023/24), benchmarked against mem, WHO-ACM and MSD.

**Results:** aedseo signalled onset in 60/63 sources and mem crossed its epidemic threshold in 55/63. When both signalled, aedseo was earlier in 45. Median lead times (weeks) were 6.5 for influenza, 4.5 for RSV, 22 for ARI and 5.5 for ILI; growth between signals was usually sustained, particularly for influenza, RSV and ILI. aedseo provided more consistent intensity categorisation across seasons than mem, WHO-ACM and MSD.

**Conclusion:** aedseo excels in early onset detection and within-season intensity assessment. It has demonstrated robust performance in the Danish national respiratory surveillance system and across European data sources, supporting timely planning and communication within surveillance frameworks.

## Introduction

During the autumn and winter seasons, multiple respiratory pathogens circulate, leading to infections and hospital admissions. The onset of circulation varies among pathogens and seasons. Likewise, the associated disease burden differs across age groups and for each pathogen [1,2,3,4]. To enable hospitals and healthcare workers to plan effectively during the respiratory disease season, robust surveillance methods are essential. Such systems should be capable of detecting increases in pathogen circulation and hospital admissions as early as possible.

Current surveillance methods to detect increase in respiratory pathogen circulation include the Moving Epidemic Method (mem [5]), the World Health Organisation (WHO) Average Curve Method (WHO-ACM [6,7,8]) and, more recently, the Mean Standard Deviation method (MSD [9]). mem and WHO-ACM classify epidemic intensity levels based on weekly peak observations from historical seasons. This approach restricts their applicability to peak-level classifications and overlooks within-season dynamics. Moreover, they require at least five historical seasons, making them less suitable when data is limited. The MSD method partially addresses this issue by using just one previous season; however, this comes at the cost of disregarding longer-term historical variability. Neither of the methods have a feature that allows for adaptability and rely on the user removing outlier seasons to get more accurate estimates for intensity levels. mem’s detection of the epidemic onset relies on thresholds derived from historical pre-epidemic periods, making it sensitive to outlier seasons, which by default remain influential for up to ten years due to mem’s long effective memory. This sensitivity complicates intensity level estimation and epidemic onset determination, especially when data quality fluctuates due to changes in e.g. surveillance methods or emergence of new subtypes.

The aim of this study was to overcome the limitations of existing methods and to develop a robust method that enables uniform assessment of infection or admission intensity levels over time, allowing comparison between seasons and early detection of increases in respiratory pathogen circulation preceding the seasonal peak. This paper introduces such a new method, the Automated and Early Detection of Seasonal Epidemic Onset and Burden Levels (aedseo [10]), which is already implemented in the Danish national respiratory surveillance system, where it has proven highly useful for weekly monitoring and communication of within-season intensity and early increase in respiratory disease activity, even when historical data is limited or fluctuating.

## Methods

### Automatic and Early detection of Seasonal Onset and Burden Levels (aedseo)

The aedseo model was developed using Danish respiratory surveillance data, including infections and admissions of influenza and RSV (seasons 2010/2011-2023/2024), and SARS-CoV-2 (seasons 2020/2021-2023/2024). The surveillance data is reported in weeks; hence we will refer it as weeks going forward.

An exponential growth rate-based method is chosen to determine the onset of seasonal waves. The objective is to identify the time point when subsequent observations are expected to keep increasing towards the seasonal peak. Because count data (observations) often exhibit overdispersion, where the variance exceeds the mean, the model employs a generalized linear model (GLM) with a quasi-Poisson family, used to model the observations as a function of time, while accounting for potential overdispersion:

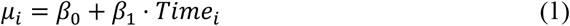

Observations (*y*_*i*_) are assumed to follow a quasi-Poisson distribution with intensity exp(*μ*_*i*_). Where *β*_0_ is the intercept and *β*_1_ is the coefficient for the *Time* predictor that represents the growth in number of new observations per time point. A rolling window, default *k* = 5 [11], is used to create subsets of the data for model fitting. This means that for each time point, the model is fitted using the number of observations from the current time point and previous *k* − 1 time points.

The growth rate, *β*_1_, is significant and positive when the 95% confidence interval lies above zero, which may signal the onset of a seasonal wave. The confidence interval is estimated using profile log-likelihood (confint function in R).

When the number of observations is low there is a risk that randomness will result in significant growth estimates in isolated periods. To improve robustness, we introduce a disease-specific threshold (*T*_*disease*_), designed to identify sustained growth indicative of a true seasonal onset. We determine *T*_*disease*_ by analysing historical periods where growth was consistently significant, focusing on periods leading to peaks in recent seasons as they best represent current trends. A rolling five-week average observation count is calculated across the entire time series. The disease-specific threshold is defined as the five-week average where sustained growth typically culminates in a seasonal peak.

Breakpoints (very low, low, medium, and high) separating intensity levels (very low, low, medium, high and very high) are utilized to monitor the intensity of weekly observations based on observations from previous seasons. Historical data from all available seasons is used to establish these breakpoints for the upcoming season. Breakpoints are determined using the three highest observations from each season, if they surpass *T*_*disease*_ to prevent seasons with very low observations to be included:

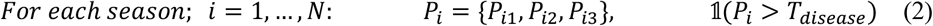

where *N* is the number of seasons and *P*_*ij*_ denotes the *j* -th highest observations in season *i*. To prevent extreme observations of a single season from disproportionately influencing the upper percentile of subsequent seasons, a log-normal distribution is employed to model the three highest observations from each previous season. This distribution aligns well with the nature of epidemic data, which often exhibits multiplicative growth patterns. The weighted negative log-likelihood, *ℒ*(*θ*), of the three highest observations from each season with an exponentially decaying weight, is minimised to estimate the parameters of the log-normal distribution:

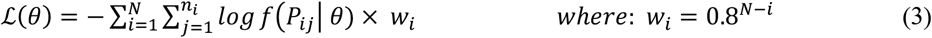

where *N* represents the total number of seasons, *n*_*i*_ is the number of observations selected from season *i* . *f*(*P*_*ij*_|*θ*) is the probability density function of the log-normal distribution with parameters, *θ* (mean and standard deviation). *w*_*i*_ = *α*^*N*−*i*^ is the weight assigned to season *i* where *α* = 0.8 is the default exponential decay factor. The decay factor was selected to balance the influence of recent and older seasons. In simple exponential smoothing, the effective memory is 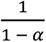 [12]. Setting *α* = 0.8 therefore gives an effective memory of about five seasons, providing enough history to smooth random year-to-year noise while remaining responsive to recent shifts.

The 97.5th percentile of the fitted distribution is used to define the high breakpoint. *T*_*disease*_ defines the very low breakpoint. The low and medium breakpoints are determined such that there is the same relative increase between each breakpoint. This is achieved by establishing logarithmic scaling between the very low and the high breakpoints. This approach maintains balanced categorization across all breakpoints, allowing meaningful intensity classification and surveillance of the current observations.

As seasonal onset is declared once the average observation count over five weeks crosses *T*_*disease*_ and is accompanied by a significant positive growth rate, the seasonal onset is typically flagged when in the low or medium intensity level.

Interpretation of observations in intensity level categories:

- Very low: Background inter-epidemic occurrence (baseline).
- Low: Above baseline, yet modest. A rapid upswing here may signal seasonal onset.
- Medium: Broadly in line with typical medium intensity.
- High: Getting closer to the upper band of this level approaches the upper tail of the historical distribution.
- Very high: Beyond the 97.5th percentile of previous peaks, i.e. getting higher than what is expected based on historical data.

### Parameter selection

Sensitivity analysis and simulation studies (Supplement S4 and S5) were conducted to:

- Select the disease-specific threshold (Fig. S4; Table S4-1).
- Choose the default sliding-window size for estimating weekly growth rates (Tables S4-1:2).
- Determine how many of the peak observations from each historical season should be selected by default when estimating intensity levels (Figs. S5-1:2).

### Benchmark

aedseo is benchmarked to already established methods; mem [5], WHO-ACM [6,7,8] and MSD [9]. mem estimates an epidemic threshold and intensity levels, which can be benchmarked with aedseos seasonal onset and intensity levels, whereas WHO-ACM and MSD methods only establish intensity levels. mem was used with default parameters from the mem model function in the mem R package. MSD was used as described in the method paper [9]. The WHO-ACM method was used with the geometric option for skewed peaks, except when producing NA values for the intensity breakpoints as it is not possible to apply the logarithm to zero observations. Then instead the standard method was used.

### Post-Onset Growth Evaluation

In the absence of a golden standard for detecting the onset of a seasonal epidemic wave, we compared growth between detections by mem (epidemic threshold) and aedseo (seasonal onset) respectively. To evaluate the accuracy of these models, it can be assumed that once a season is detected, there should be continuous growth in observations in the following weeks. If growth does not follow the detected onset, the detection may be considered a false positive, indicating that the season likely began later than identified. The evaluation is conducted by extracting the time between the detection for both models and assessing whether significant growth occurs or not. This comparison allows for performance evaluation of both models in early detection of seasonal epidemic waves.

### Validation/test of model using various European data sources

The European Respiratory Virus Surveillance Summary (ERVISS [13]), a joint initiative of ECDC and the WHO Regional Office for Europe, provides weekly aggregated data sets of respiratory viral infections by country in the European Union, collected based on various surveillance systems. The following types of surveillance data sources were downloaded from ECDC’s GitHub [14] to validate our aedseo method:

#### Sentinel syndromic data

Observations of influenza-like illness (ILI) and acute respiratory infection (ARI) that rely on syndromic case definitions. In most countries, ILI/ARI surveillance is conducted by sentinel networks of primary-care practices, intended to be nationally representative, though coverage may vary. In some countries, surveillance is universal, covering the entire population [13].

#### Non-sentinel laboratory data

National diagnostic and reference laboratories report weekly totals of laboratory-confirmed detections of influenza-virus and RSV irrespective of the patient’s clinical presentation or testing venue (hospital, outpatient clinic, etc.) [13].

For both data sources, countries report the ISO-week number and the corresponding number of observations or detections (collectively referred to as observations throughout this article). We define the surveillance year from week 21 in one year to week 20 in the following calendar year to ensure that complete epidemic cycles are captured, accommodating pathogens whose peaks fall earlier or later in the season. There is currently data available from season 2014/2015 to season 2024/2025 for a majority of European countries. Data sources included in the study have reported at least five seasons of data, have data for both influenza and RSV, have an average of minimum 15 weeks of data per season and include data for season 2024/2025. The 21 countries that satisfy the criteria are anonymized with alphabetic letters from A to U and constitute 63 data sources (Supplement S1). ILI and ARI incidence is reported per 100,000 population, except for country N and O that report per 100 consultations (and are excluded as historical consultation rates are not publicly available). For the exponential growth rate-based method ILI and ARI incidence are transformed to national population level with population data from each country (2014-2025), which is available from Eurostat [15].

## Results

Detailed results are shown for four different European countries:

- Non-sentinel influenza infections in country A.
- Non-sentinel RSV infections in country B.
- Sentinel Acute respiratory infection (ARI) in country C.
- Sentinel Influenza-like illness (ILI) in country D.

Figure 1 visualises the raw weekly observations for influenza (Country A), RSV (Country B), ARI (Country C) and ILI (Country D). Influenza and RSV show a clear seasonal pattern, with peaks generally occurring in the winter months, and the intensity of these peaks increased after the 2020/2021 season. Notably, the 2020/2021 season displays almost no detectable activity in influenza and RSV, likely due to the SARS-COV-2 pandemic and associated non-pharmaceutical interventions [16, 17].

**Figure 1.**
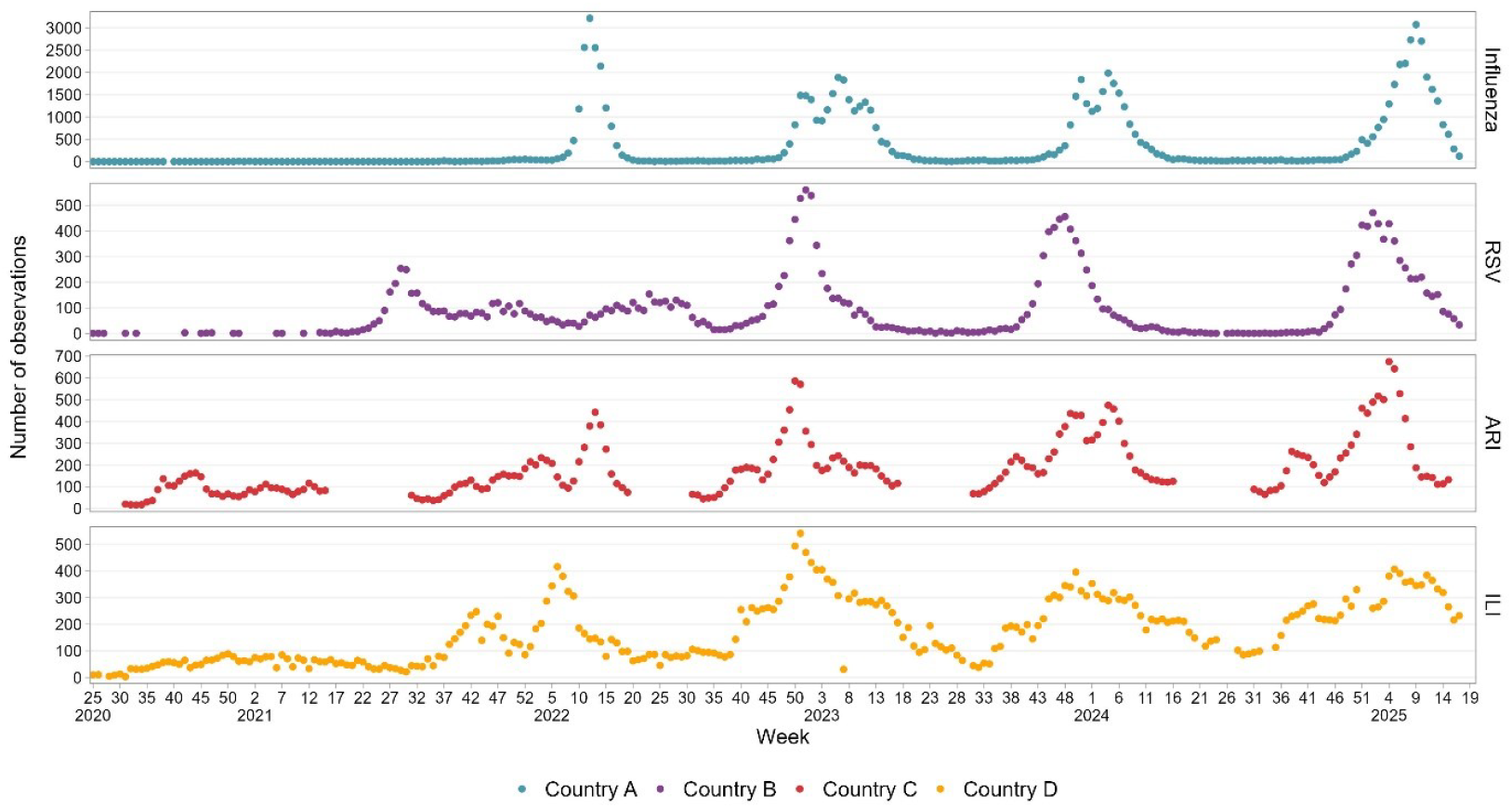
Time series of raw weekly observations for influenza (Country A), RSV (Country B), ARI (Country C) and ILI (Country D) from season 2020/2021 to 2024/2025.

### Estimating disease-specific threshold

The first step in the aedseo method is to determine the disease-specific threshold. Applying the aedseo method to our time series data excluding season 2024/2025 (Fig. 1), we obtain the following *T*_*disease*_:

- Influenza (Country A): 42.4 laboratory confirmed detections
- RSV (Country B): 10.8 laboratory confirmed detections
- ARI (Country C): 62.2 observations per 100,000 population
- ILI (Country D): 126.1 observations per 100,000 population

The disease-specific thresholds were estimated with the *estimate_disease_threshold* function from the aedseo R package which is built on the method described in the methods section. aedseo *T*_*disease*_ estimates and mem epidemic thresholds for all 63 surveillance data sources by country for season 2024/2025 are available in Supplement S2.

### Benchmark

We applied the four surveillance methods; aedseo, mem, WHO-ACM and MSD to the 63 data sources from the 21 European countries.

Seasonal onset and intensity levels for countries A-D are presented in the following sections, while results for all 21 countries including 63 data sources (21 influenza, 21 RSV, 8 ARI, 13 ILI) analysis are available in Supplement S1. Complete available time series data (Supplement S1) was used with aedseo, mem and WHO-ACM, excluding season 2020/2021 for mem for better performance. MSD was used with the previous season to the most recent season.

Figure 2 and 3 show intensity levels and seasonal onset/epidemic threshold (aedseo, mem) for the four data sources estimated by the four surveillance methods, with observations on the y-axis and weeks on the x-axis for season 2024/2025. Intensity level estimates differ markedly across methods (Figs. 2–3; Table 1). For Influenza and RSV, especially the low, medium and high (MSD and mem) and the medium and high (WHO-ACM) intensity levels are very compressed, due to the very steep peaks in previous seasons (Fig. 1). As a result, the number of weekly detections has to increase substantially to move from very low to low. For WHO-ACM the low intensity level is very broad, due to the very low breakpoint as it reflects the mean of the peak observations from previous seasons (Fig. 1). The two most recent influenza seasons reach approximately 2000 observations, while the third most recent reaches 3214 observations, which is slightly higher than season 2024/2025 on 3071 observations (Table 1). The aedseo method marks season 2024/2025 as high level for a long period but does not reach the very high level. MSD and WHO-ACM on the other hand both reaches very high level, where mem only slightly reaches the high level.

**Table 1.** Includes information about the historical highest peak from each surveillance data source, the peak observation of season 2024/2025 and the corresponding intensity levels estimated by each method.

| Data source | Historical highest peak observation | Peak observation season 2024/2025 | Intensity levels | aedseo | MSD | mem | WHO-ACM |
| --- | --- | --- | --- | --- | --- | --- | --- |
| Influenza (Country A) | 2021/2022<br>3214 | 3071 | very low<br>low<br>medium<br>high | 42.4<br>190.4<br>845.9<br>3838.7 | 213.5<br>672.1<br>1589.2<br>2506.3 | 356.5<br>891<br>2718.8<br>4451.8 | 14.5<br>806.9<br>1493.9<br>2420.3 |
| RSV (Country B) | 2022/2023<br>560 | 471 | very low<br>low<br>medium<br>high | 10.8<br>44<br>178.5<br>724.2 | 62.7<br>149.9<br>324.2<br>498.5 | 110.5<br>187.6<br>405<br>569.2 | 30<br>178.8<br>269.7<br>372.1 |
| ARI (Country C) | 2022/2023<br>586.2 | 675 | very low<br>low<br>medium<br>high | 62.2<br>145<br>338<br>787.8 | 167.6<br>283.2<br>514.2<br>745.3 | 242.1<br>349.4<br>515.4<br>612 | 137.6<br>407.7<br>600.3<br>931 |
| ILI (Country D) | 2022/2023<br>541.3 | 406.8 | very low<br>low<br>medium<br>high | 126.1<br>217.3<br>374.7<br>646 | 103.7<br>195.7<br>379.4<br>563.3 | 259.8<br>207.8<br>413.1<br>559.8 | 75.9<br>262.6<br>313.3<br>360.9 |

**Figure 2.**
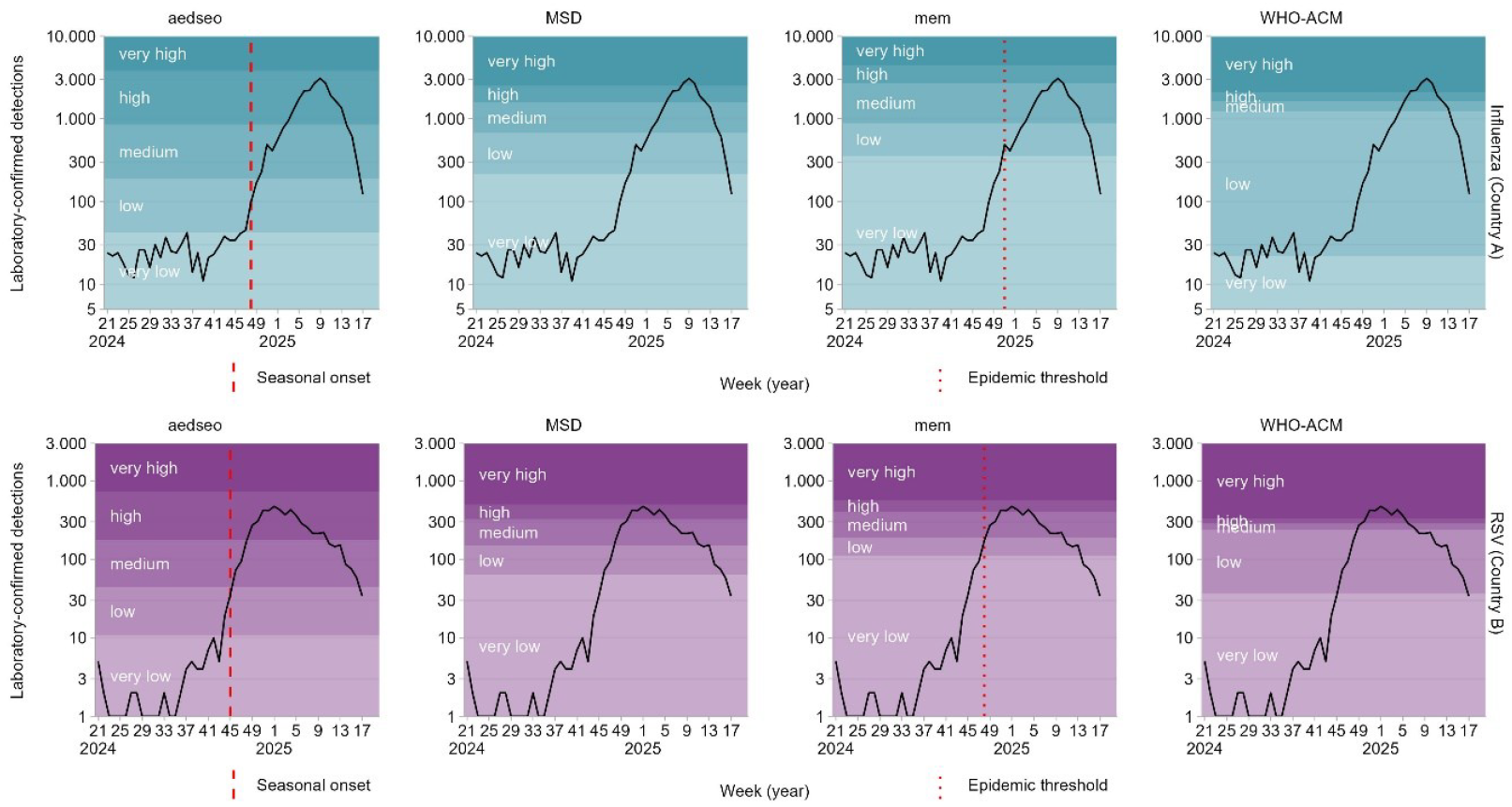
Comparing the four different surveillance methods on non-sentinel laboratory data: Weekly totals of laboratory-confirmed detections of influenza-virus (Country A) top row and RSV (Country B) bottom row, 2024/2025.

**Figure 3.**
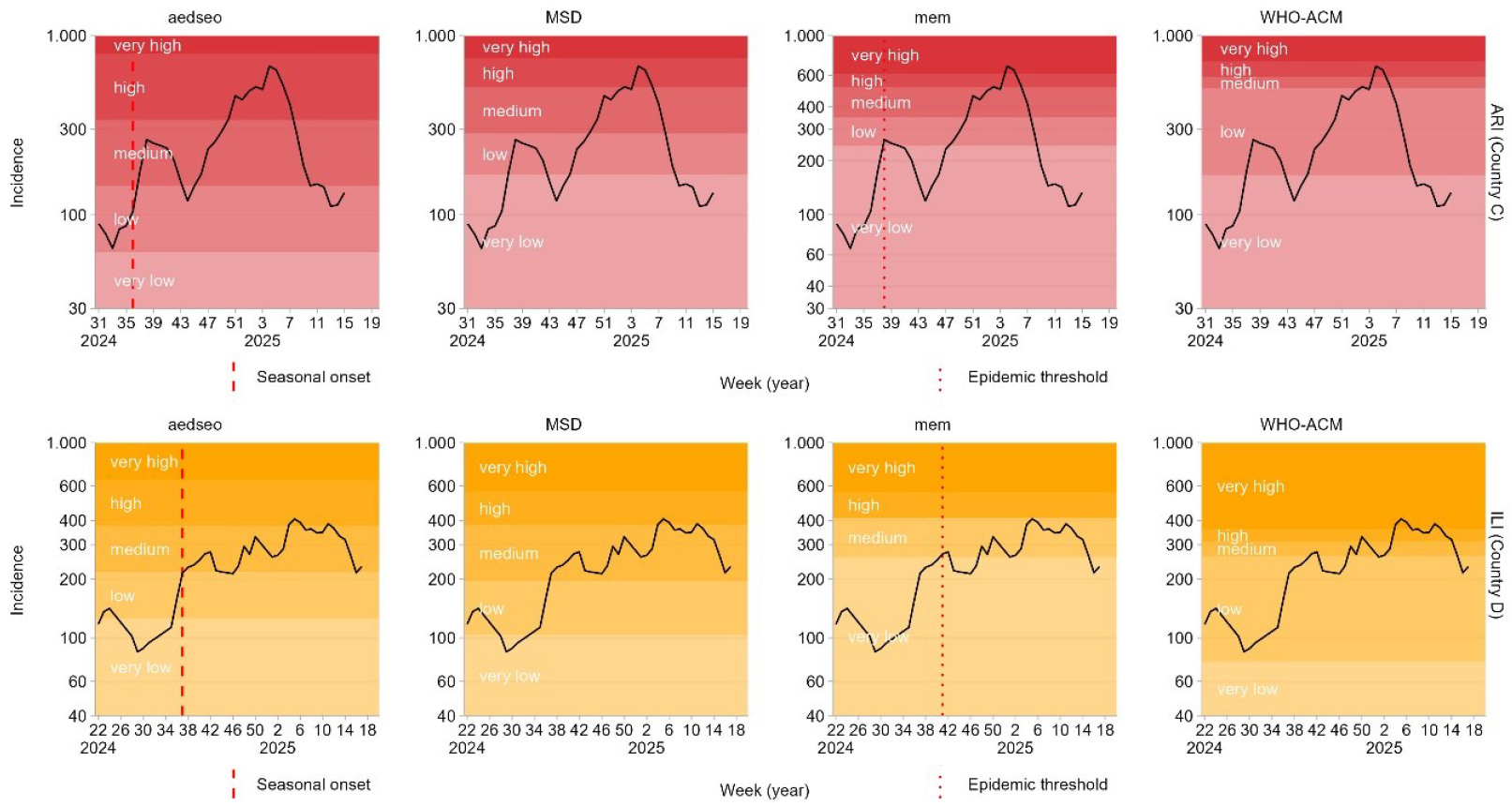
Comparing the four different surveillance methods on sentinel syndromic data: Weekly incidence of influenza-like illness (Country C) top row and acute respiratory infection (Country D) bottom row that rely on syndromic case definitions, season 2024/2025.

ILI and ARI represent symptom-based syndromes caused by multiple pathogens, so the activity can fluctuate through the surveillance year, and even create multiple peaks. However, figure 1 shows that their highest peaks coincide with the period when influenza and RSV have their peaks for the corresponding countries C and D (Supplement S1). Outside that window (influenza and RSV), activity is likely driven by pathogens with weaker or a different seasonality.

For ILI (Country D; Fig. 3; Table 1), mem does not have a low intensity level. The reason can be seen in Table 1 where the very low breakpoint (the epidemic threshold) exceeds the low breakpoint, hence skipping the low intensity level. The same can be observed for countries A (RSV), D (ILI), E (ARI, influenza), F (RSV), H (influenza, RSV), J (RSV), K (influenza, RSV), L (ILI), M (ARI), N (influenza, RSV), Q (RSV), R (RSV) and S (ARI) (Supplement S1).

A seasonal onset was detected in 60 out of 63 of the data sources by aedseo and the epidemic threshold was reached in 55 out of 63 data sources using mem. Out of these 55 data sources aedseo detected the seasonal onset earlier than mem’s epidemic threshold 45 times, whereas mem detected it earlier 7 times, and in the remaining 3 data sources it was detected in the same week (Supplement S3).

Table 2 shows the median and IQR for the number of weeks between the seasonal onset for aedseo and the epidemic threshold for mem, it includes 45 data sources for season 2024/2025 where both methods had a detection. For influenza and RSV seasonal onset is detected 6.5 (3-12.25) and 4.5 (4-12.25) weeks before mem reaches the epidemic threshold. The majority of these intermediate weeks has significant growth. The 7 data sources (3 Influenza, 3 RSV, 1 ILI) where mem detected seasonal onset before aedseo, have a median of 1 (influenza and RSV) and 2 (ILI) intermediate weeks between detections (Supplement S3; Table S3-2). For ARI and ILI aedseo detected the season 22 (11-29) and 5.5 (3-14.25) weeks prior to mem, respectively. For ARI the intermediate weeks have a very large range of non-significant growth, whereas ILI shows significant growth in majority of the intermediate weeks.

**Table 2.** Median and IQR of the number of weeks between aedseo seasonal onset and mem’s epidemic threshold for the 45 data sources (Table S3-1) where aedseo detected seasonal onset prior to mem’s epidemic threshold.

| **Data source** | **No. of data sources with seasonal onset before mem epi. threshold** | **median IQR**<br>**No. of weeks between seasonal onset vs. mem epi. threshold** | **median IQR significant growth weeks** | **median IQR no significant growth weeks** |
| --- | --- | --- | --- | --- |
| Influenza | 16 | 6.5 (3 - 12.25) | 5 (2.75 - 7) | 1 (0 - 5) |
| RSV | 14 | 4.5 (4 - 12.25) | 4.5 (4 - 7.75) | 0 (0 - 4.5) |
| ARI | 5 | 22 (11 - 29) | 8 (7 - 8) | 14 (4 - 20) |
| ILI | 10 | 5.5 (3 - 14.25) | 4 (3 - 6) | 1.5 (0 - 4.75) |

## Discussion

Based on Danish respiratory surveillance data we developed a method that provides robust, disease-specific assessments of infection and hospital admission intensity levels, allowing for within-season intensity assessment based on historical data. In addition, the method can detect early increases in circulating respiratory pathogens, which is crucial for informing healthcare professionals and enabling proactive planning. The approach has also proven valuable for consistent public and media messaging of epidemiological surveillance data. Applied to diverse European datasets, the method has demonstrated its ability to generate reliable intensity level estimates both across pathogens, countries and for different types of surveillance sources.

As ILI and ARI can fluctuate a lot and have multiple peaks (Supplement S1), estimating seasonal onset might not be appropriate. Instead we suggest to track the intensity level during the season and only use the aedseo seasonal onset method for the disease-specific analysis. If the desire is to estimate multiple onsets, the *combined_seasonal_output* function in the aedseo package can also be used for this purpose.

In practice ILI and ARI should be comparable between countries, but due to differences in data collection this might not always be the case. In contrast, non-sentinel data for influenza and RSV is even harder to compare internationally because the scale, geographic coverage, data quality and testing criteria and volume differ from one country to another, even between seasons. Accordingly, the aedseo method excels when measuring disease intensity within a country for each specific pathogen (e.g. influenza, RSV and SARS-CoV-2).

The compared methods answer different questions; aedseo supports within-season intensity assessment and indicates when observations cross the 97.5th percentile derived from weighted historical peaks (Figs. 2 – 3; Supplement S1), whereas mem and WHO-ACM are primarily retrospective, placing the final peak within the 40th/90th/97.5th percentiles of peaks from previous seasons (Fig. 2 – 3; Supplement S1), which is valuable for post-season benchmarking. In 17/63 data sources mem’s epidemic threshold is above the low breakpoint, emphasising why a disease-specific threshold is important due to inter-season variation. For readers who still want peak to peak comparisons, aedseo provides those as well within the aedseo R package.

Our findings revealed substantial inter-season variation in peak intensity in most surveillance sources (Fig. 1; Supplement S1). Such variability poses challenges for standard surveillance methods (mem, WHO-ACM), which rely on assumptions of minimal fluctuation in the data and thus struggle under significant inter-seasonal variation (Figs. 2-3; Supplement S1). This highlights the importance of employing a robust tool, such as aedseo, that adaptively manages inter-season variations without the need to exclude seasons. This adaptive capacity is particularly crucial when historical data are limited or atypical, e.g. due to changes in testing strategies, or during pandemic years.

A limitation of all the methods is that a season with an extreme peak (e.g. with unusual increased testing) can drastically increase the intensity levels for the next season. If subsequent seasons then revert to previous patterns, mem by default will “remember” such an extreme season for up to ten seasons going forward, whereas aedseo can adjust quickly due to the decay factor in the model. This parameter can be tuned to increase or decrease the influence of older seasons when using the model if needed.

The high breakpoint (between the high and very high burden levels) estimation differs mainly in how many peak observations each method uses, it decreases with more observations and vice versa. As we want the high breakpoint to be exceeded only in truly extreme seasons, we select the three peak observations from each season in the aedseo method (Supplement S5). mem uses 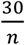 where *n* is the number of seasons, hence it uses more than three peak observations when there are fewer than ten seasons, causing the high breakpoint to shift downward (Supplement S5). WHO-ACM uses one peak from each season making it more vulnerable to random variation. MSD does not use only peak observations, but the mean and standard deviation of the entire previous season.

The aedseo model was also applied to SARI surveillance data but analysis was excluded as SARI data is reported for catchment populations which are not publicly available on erviss [13,14]. Additionally, SARS-CoV-2 was excluded as only aedseo and MSD can be used for this, due to the limited historical data (one season) available post–mass-testing.

## Conclusion

The proposed aedseo method has shown robust early warning of circulating respiratory disease activity and reliable monitoring of within-season intensity across data sources/countries compared with current methods. It is integrated in Denmark, where it has proven highly useful in real-world surveillance settings and consistent public health communication during and between respiratory seasons.

## Data Availability

All data produced are available online at https://github.com/EU-ECDC/Respiratory_viruses_weekly_data/tree/main/data

https://ec.europa.eu/eurostat/web/population-demography/demography-population-stock-balance/database

## Notes

### Competing Interest Statement

The authors have declared no competing interest.

### Clinical Protocols

https://github.com/ssi-dk/aedseo

### Funding Statement

This study did not receive any funding.
We thank Luisa Hallmaier-Wacker, Expert respiratory viruses and Nick Bundle, Principal
expert respiratory viruses, from the European Centre for Disease Prevention and Control
(ECDC) for their valuable advice regarding interpretation of the respiratory virus
surveillance data from the ECDC github.

### Author Declarations

This study is based entirely on publicly available, aggregated surveillance data from the European Centre for Disease Prevention and Control (ECDC) and the World Health Organisation Regional Office for Europe (ERVISS platform). No individual-level or identifiable data were used; therefore, ethical approval and informed consent were not required. ECDC Github repository: https://github.com/EU-ECDC/Respiratory_viruses_weekly_data/tree/main/data

