## Supplementary Material for "A method for robust estimation of seasonal onset and intensity in respiratory surveillance data: evaluated using data from 21 European countries"

This supplementary material should be used alongside the article *"A method for robust estimation of seasonal onset and intensity in respiratory surveillance data: evaluated using data from 21 European countries"*, on behalf of the authors, who remain responsible for the accuracy and appropriateness of the content. The same standards for ethics, copyright, attributions and permissions as for the article apply.

**Supplement S1 (pages 2-22):** Time series plots and onset and intensity plots for countries A-U, season 2024/2025 with methods (aedseo, MSD, mem, WHO-AMD) from the corresponding available seasons of each data source.

**Supplement S2 (pages 23-24):** All disease-specific thresholds (aedseo) and epidemic thresholds (mem) for countries A-U, season 2024/2025. Influenza and RSV are reported as laboratory-confirmed detections. ARI and ILI are reported as observations per 100,000 population.

**Supplement S3 (pages 25-27):** Seasonal onset/epidemic threshold timing estimated by the aedseo and mem methods of season 2024/2025 for the selected 21 countries and 63 data sources. Countries D (RSV) and M (influenza and RSV) are not included as they did not reach a signal in any of the methods. Countries D (influenza, ARI), E (ARI), K (ILI) and M (ARI) are not included as it was only aedseo that had a signal in these data sources.

**Supplement: S4 (pages 28-29)**: Sensitivity analysis of the disease-specific threshold and sliding window size.

**Supplement S5 (page 30):** Sensitivity analysis of number of peak observations for intensity level estimates.

### Supplement S1

#### Country A

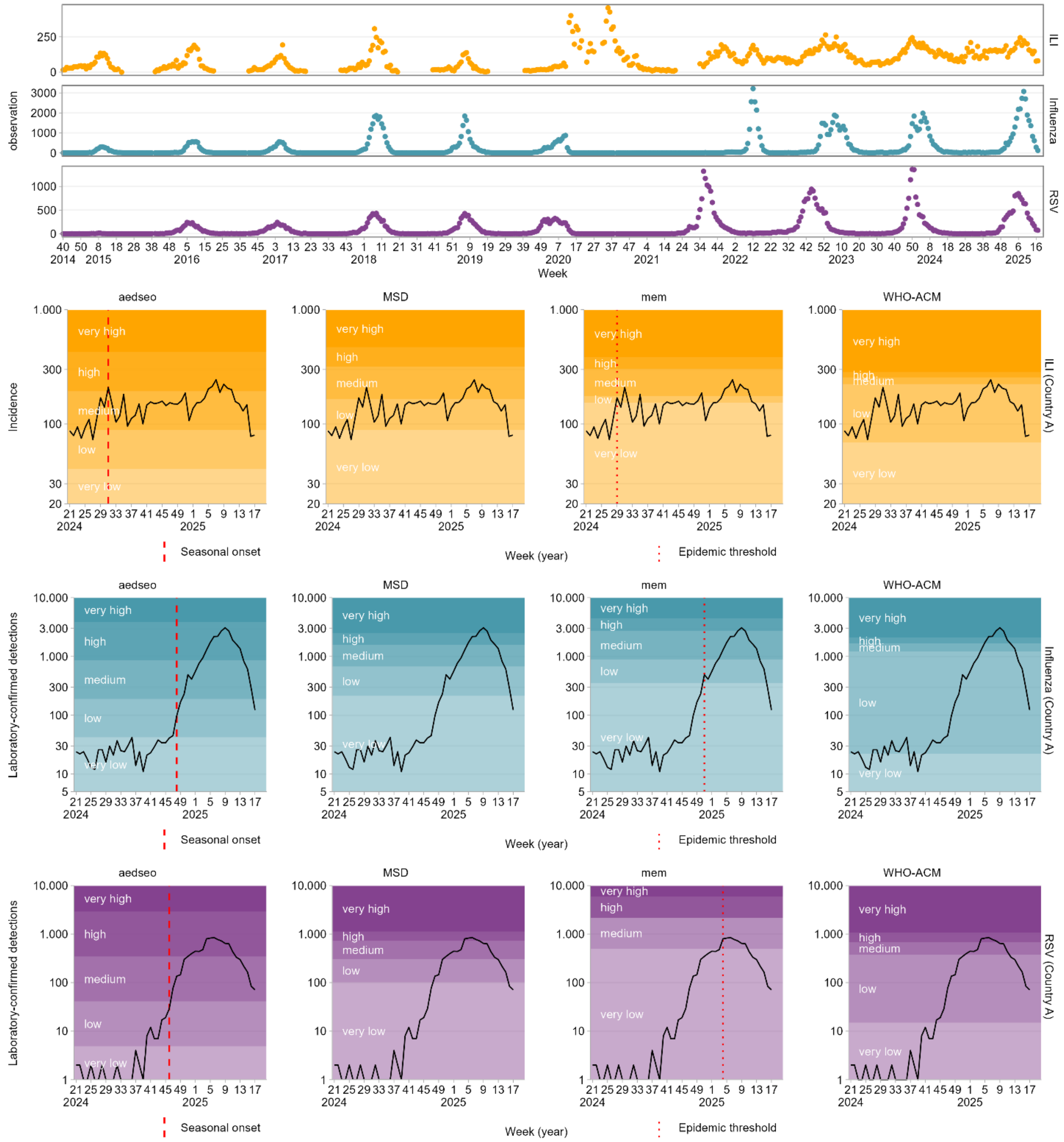

#### Country B

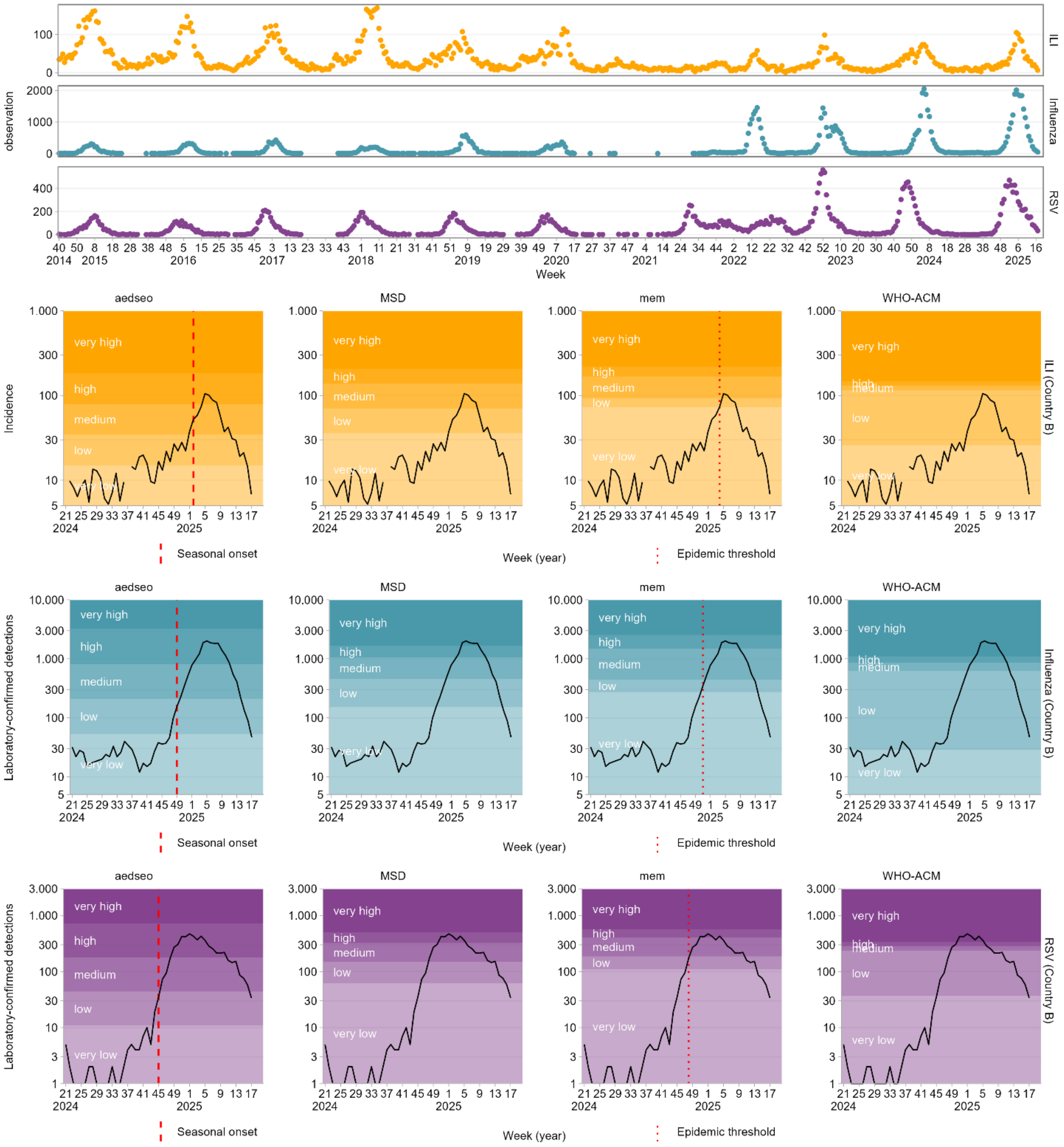
ILI
Influenza
RSV
observation
Week
aedseo
MSD
mem
WHO-ACM
Incidence
ILI (Country B)
Influenza (Country B)
RSV (Country B)
Laboratory-confirmed detections
very high
high
medium
low
very low
Week (year)
Seasonal onset
Epidemic threshold

### Country C

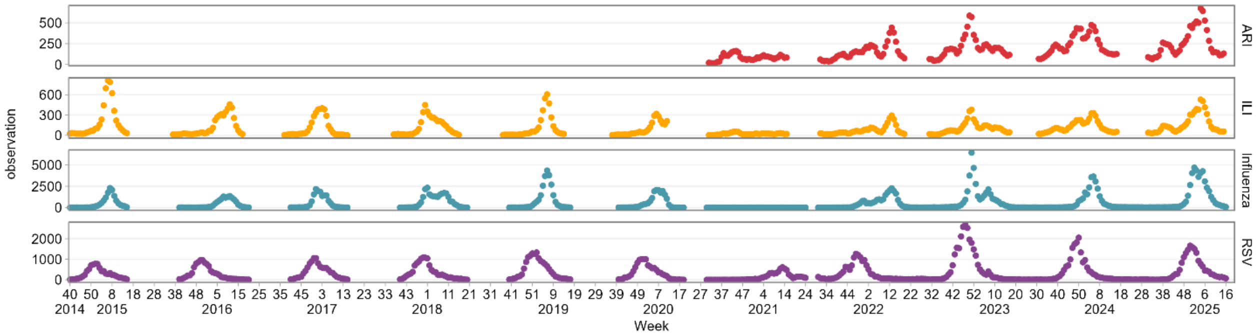

observation
ARI
ILI
Influenza
RSV
Week
2014
2015
2016
2017
2018
2019
2020
2021
2022
2023
2024
2025


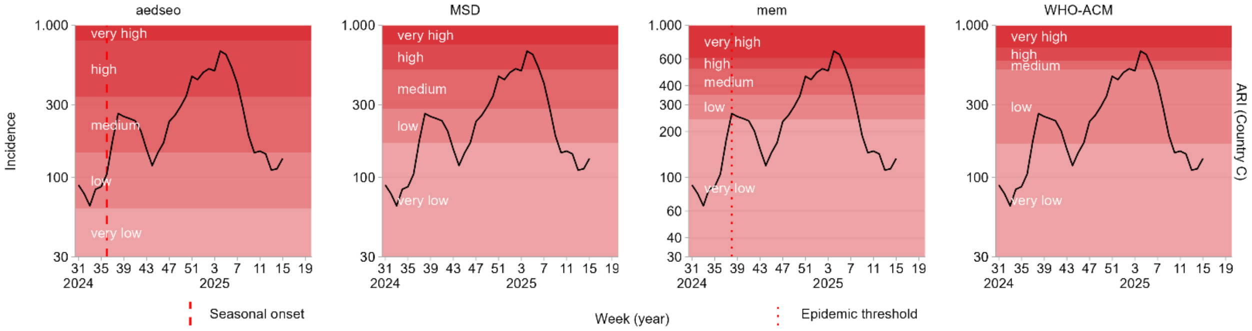

aedseo
MSD
mem
WHO-ACM
Incidence
very high
high
medium
low
very low
ARI (Country C)
Week (year)
Seasonal onset
Epidemic threshold


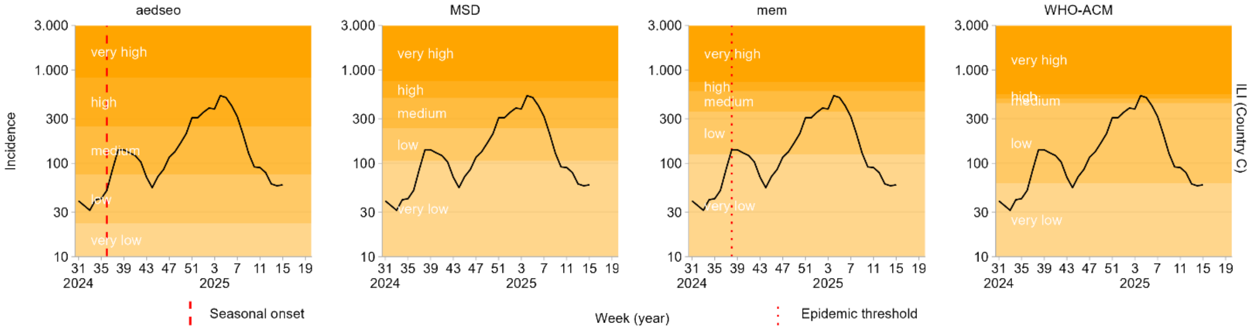

aedseo
MSD
mem
WHO-ACM
Incidence
very high
high
medium
low
very low
ILI (Country C)
Week (year)
Seasonal onset
Epidemic threshold


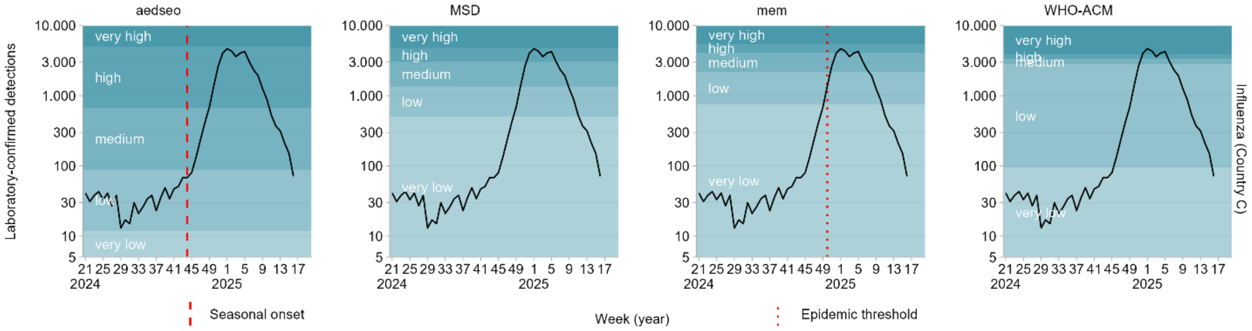

aedseo
MSD
mem
WHO-ACM
Laboratory-confirmed detections
very high
high
medium
low
very low
Influenza (Country C)
Week (year)
Seasonal onset
Epidemic threshold


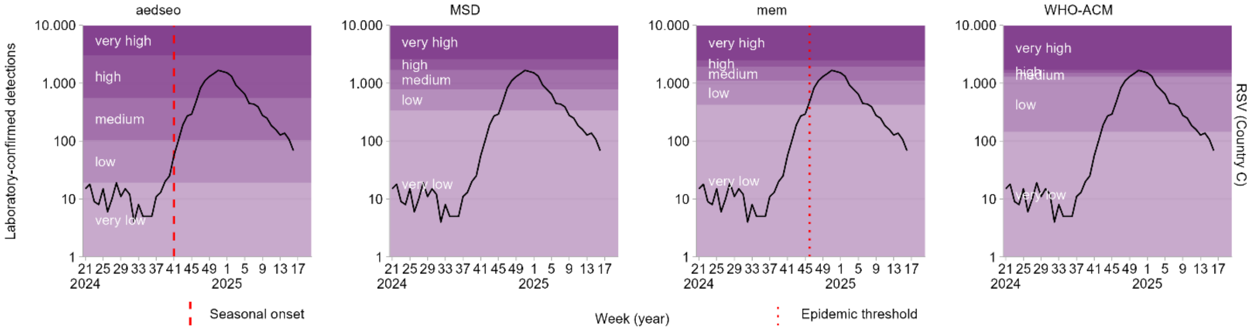

aedseo
MSD
mem
WHO-ACM
Laboratory-confirmed detections
very high
high
medium
low
very low
RSV (Country C)
Week (year)
Seasonal onset
Epidemic threshold

### Country D

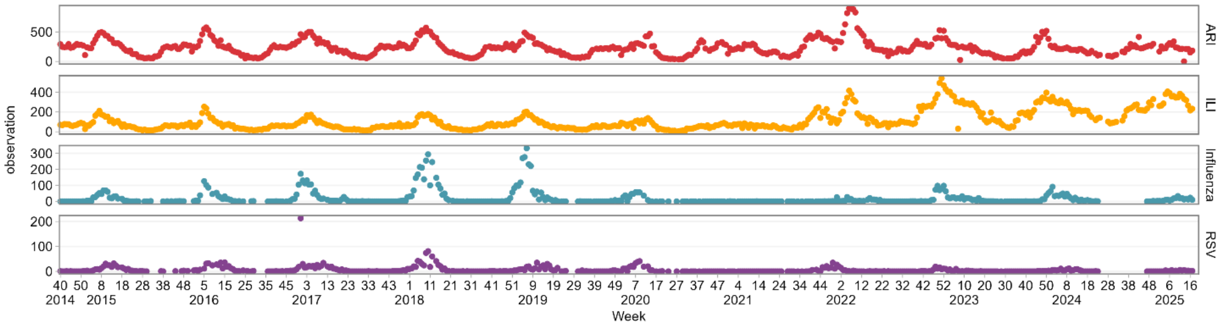
ARI
ILI
Influenza
RSV
observation
Week
2014 2015 2016 2017 2018 2019 2020 2021 2022 2023 2024 2025

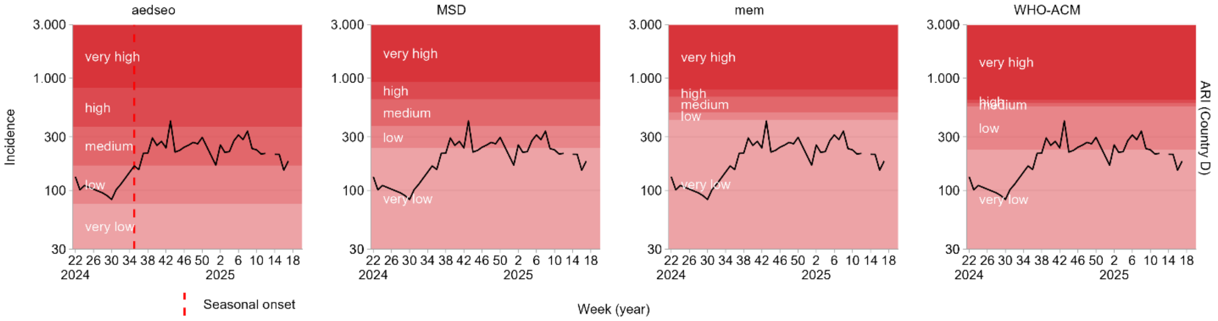
aedseo
MSD
mem
WHO-ACM
Incidence
very high
high
medium
low
very low
ARI (Country D)
Seasonal onset
Week (year)

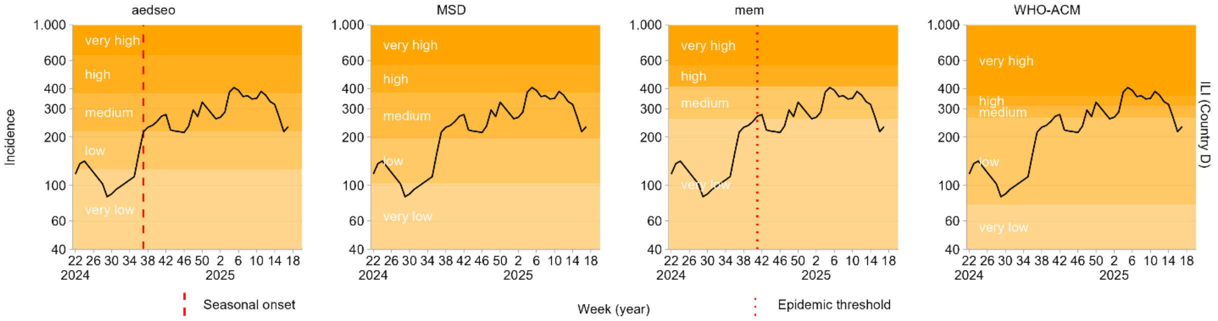
aedseo
MSD
mem
WHO-ACM
Incidence
very high
high
medium
low
very low
ILI (Country D)
Seasonal onset
Week (year)
Epidemic threshold

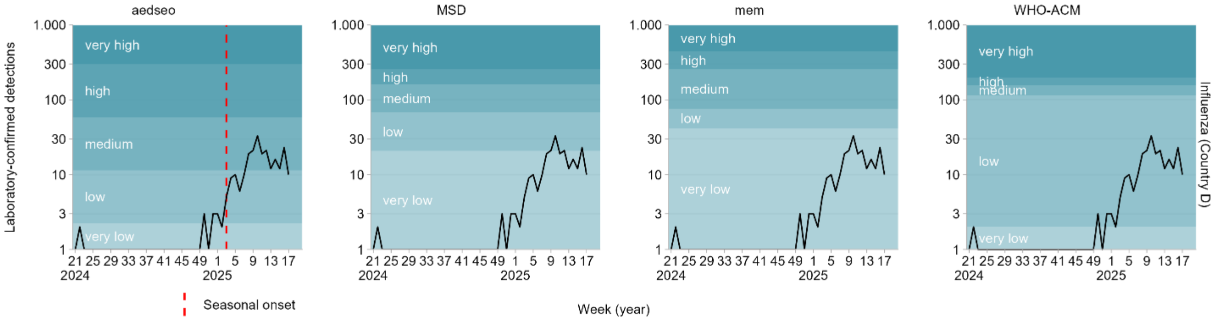
aedseo
MSD
mem
WHO-ACM
Laboratory-confirmed detections
very high
high
medium
low
very low
Influenza (Country D)
Seasonal onset
Week (year)

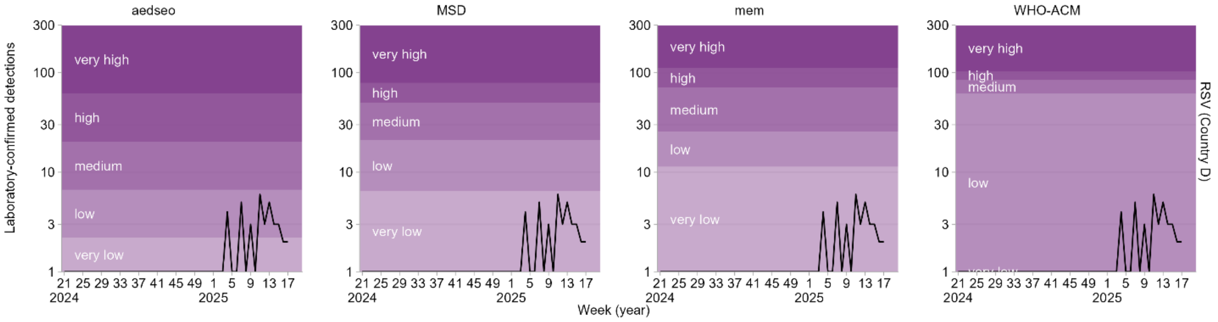
aedseo
MSD
mem
WHO-ACM
Laboratory-confirmed detections
very high
high
medium
low
very low
RSV (Country D)
Week (year)

#### Country E

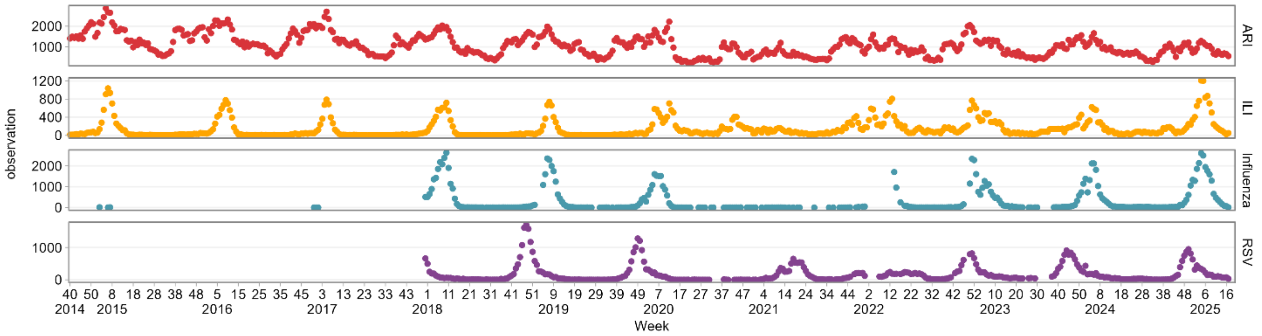

ARI
ILI
Influenza
RSV
observation
Week


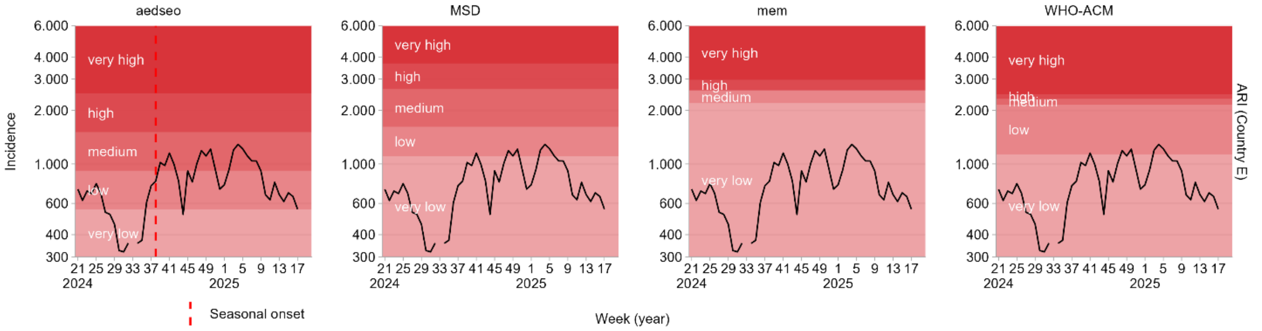

aedseo
MSD
mem
WHO-ACM
Incidence
ARI (Country E)
very high
high
medium
low
very low
Seasonal onset
Week (year)


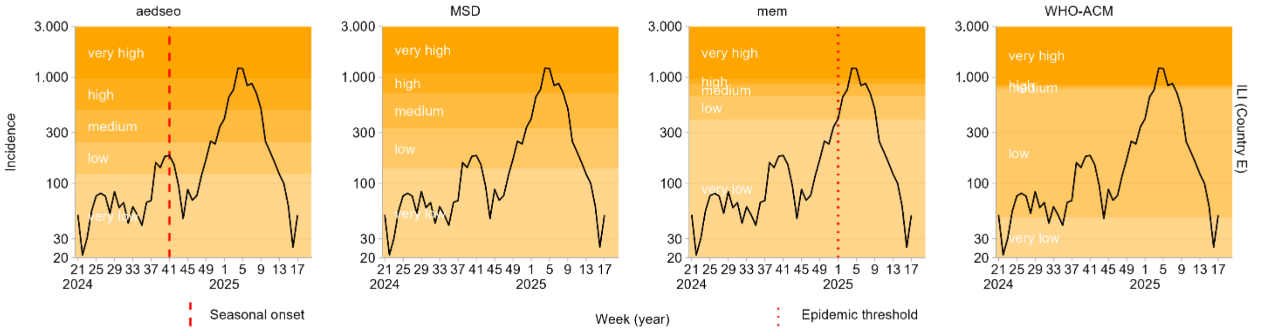

aedseo
MSD
mem
WHO-ACM
Incidence
ILI (Country E)
very high
high
medium
low
very low
Seasonal onset
Week (year)
Epidemic threshold


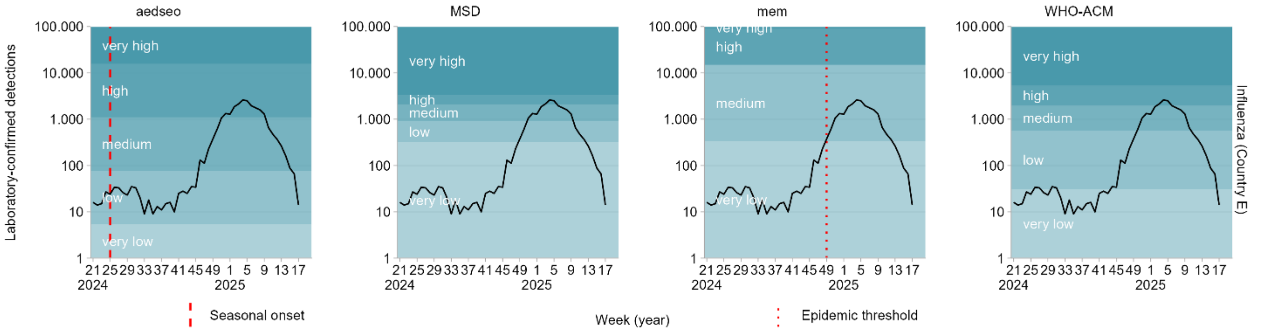

aedseo
MSD
mem
WHO-ACM
Laboratory-confirmed detections
Influenza (Country E)
very high
high
medium
low
very low
Seasonal onset
Week (year)
Epidemic threshold


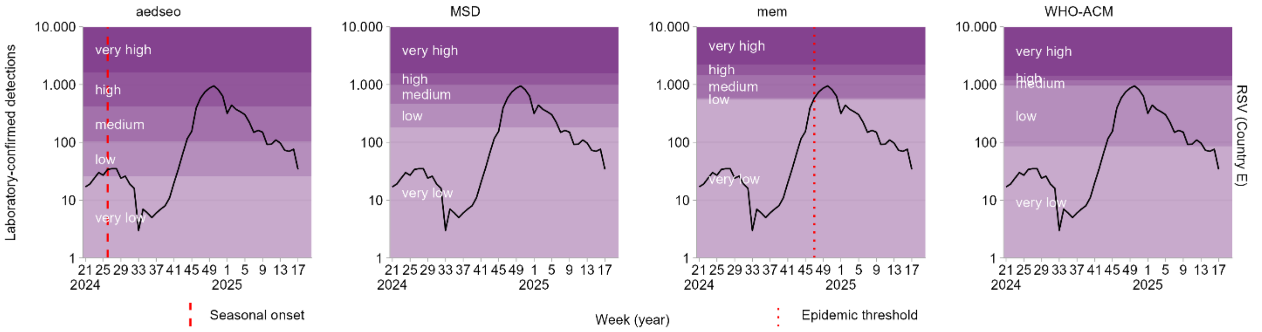

aedseo
MSD
mem
WHO-ACM
Laboratory-confirmed detections
RSV (Country E)
very high
high
medium
low
very low
Seasonal onset
Week (year)
Epidemic threshold

#### Country F

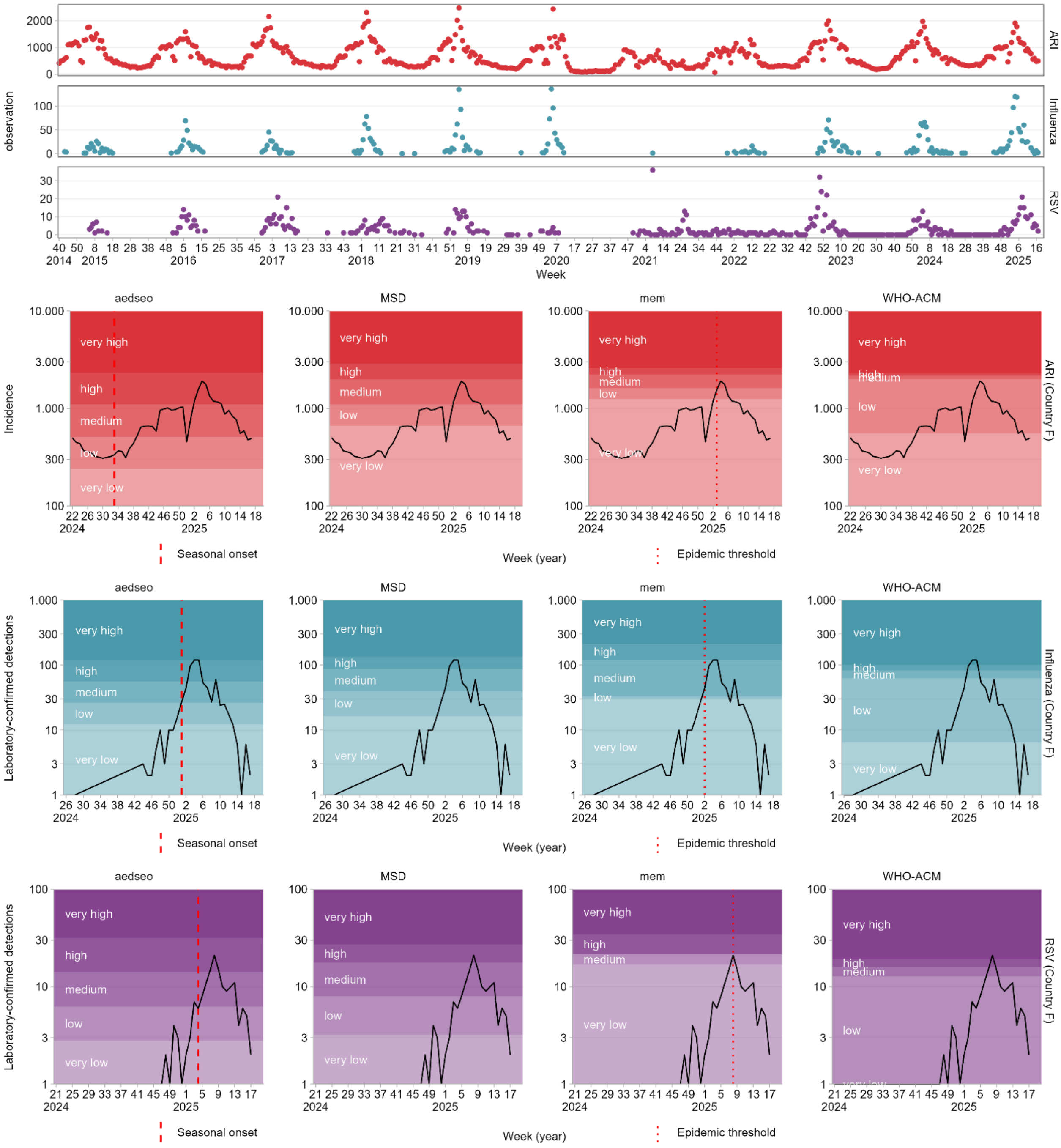

ARI
Influenza
RSV
observation
Week
aedseo
MSD
mem
WHO-ACM
very high
high
medium
low
very low
Incidence
ARI (Country F)
Laboratory-confirmed detections
Influenza (Country F)
RSV (Country F)
Week (year)
Seasonal onset
Epidemic threshold

#### Country G

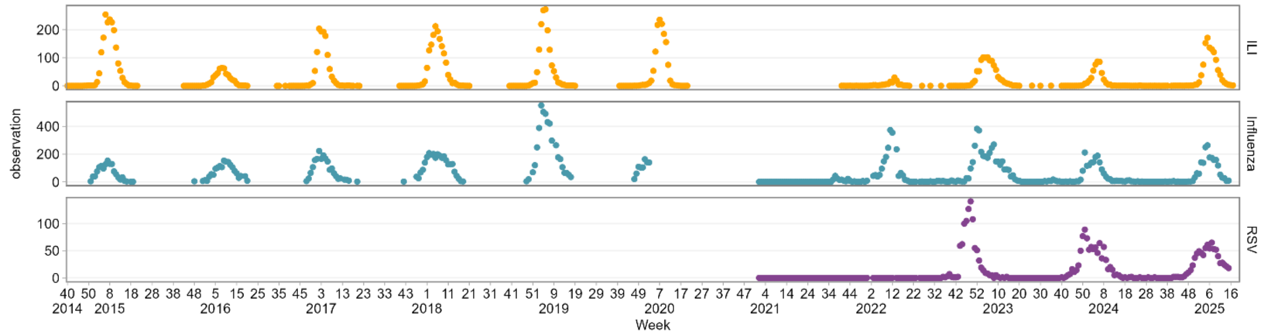

ILI
Influenza
RSV
observation
Week
2014 2015 2016 2017 2018 2019 2020 2021 2022 2023 2024 2025


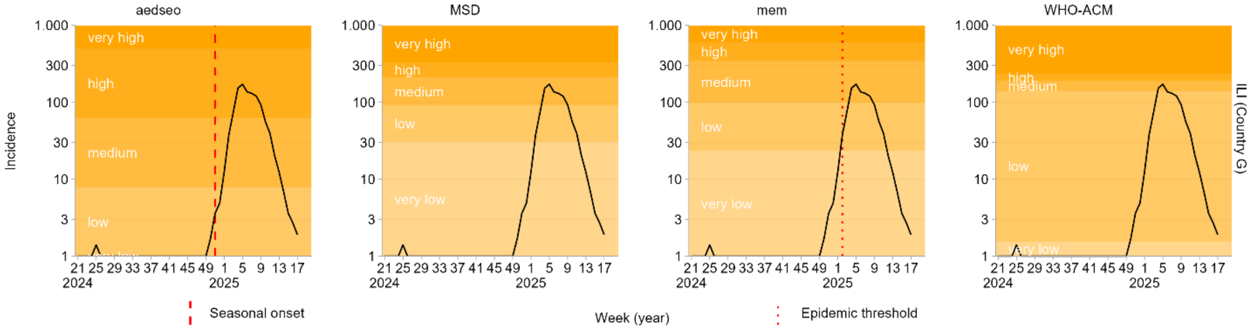

aedseo
MSD
mem
WHO-ACM
Incidence
ILI (Country G)
very high
high
medium
low
very low
Week (year)
Seasonal onset
Epidemic threshold


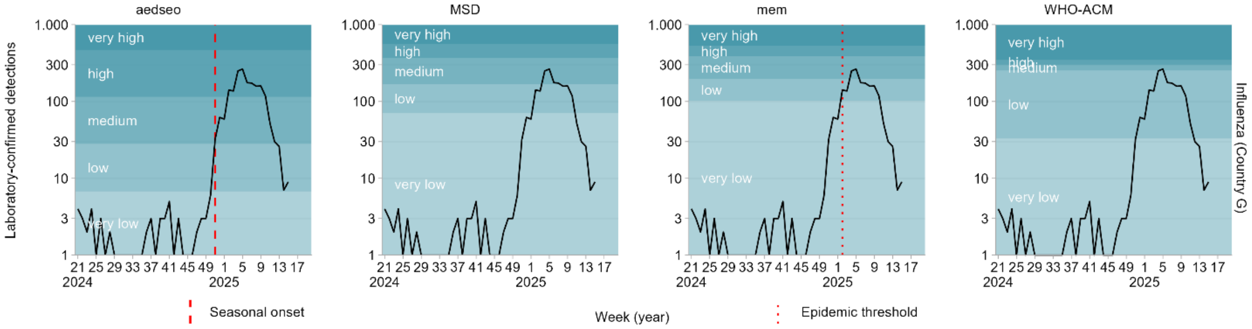

aedseo
MSD
mem
WHO-ACM
Laboratory-confirmed detections
Influenza (Country G)
very high
high
medium
low
very low
Week (year)
Seasonal onset
Epidemic threshold


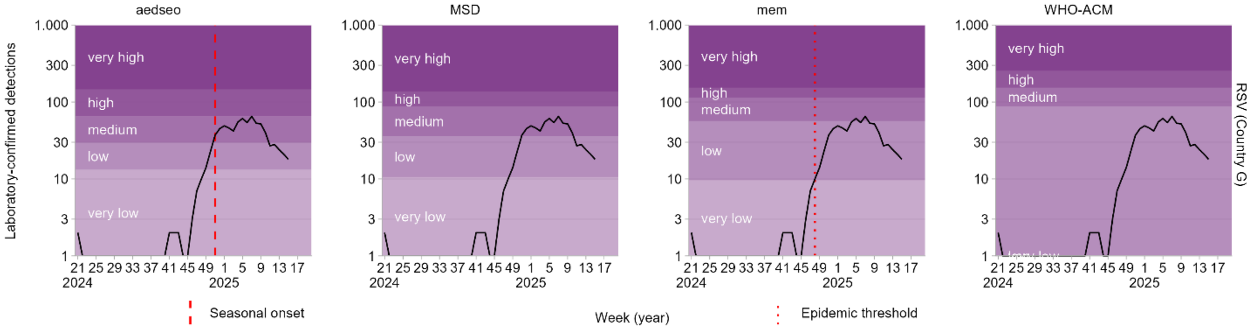

aedseo
MSD
mem
WHO-ACM
Laboratory-confirmed detections
RSV (Country G)
very high
high
medium
low
very low
Week (year)
Seasonal onset
Epidemic threshold

### Country H

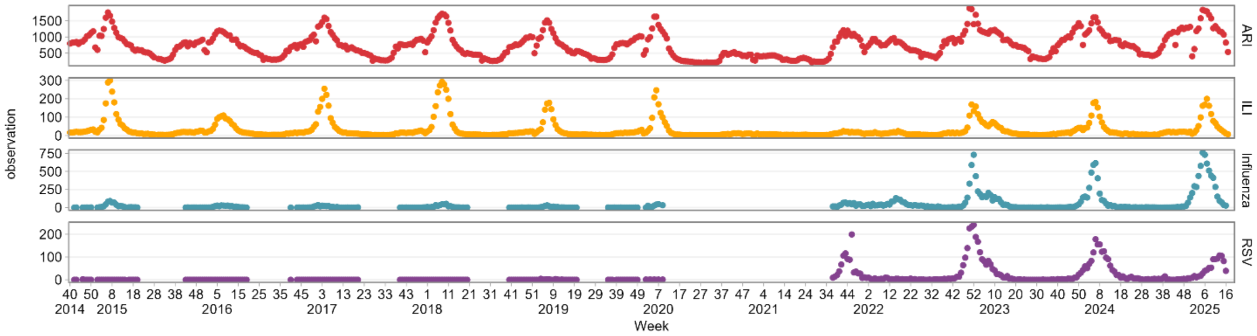
ARI
ILI
Influenza
RSV
observation
Week

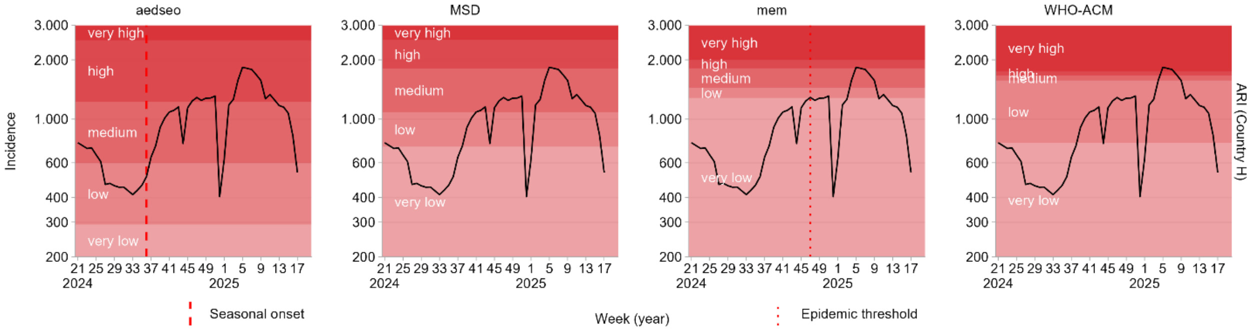
aedseo
MSD
mem
WHO-ACM
Incidence
ARI (Country H)
Seasonal onset
Week (year)
Epidemic threshold

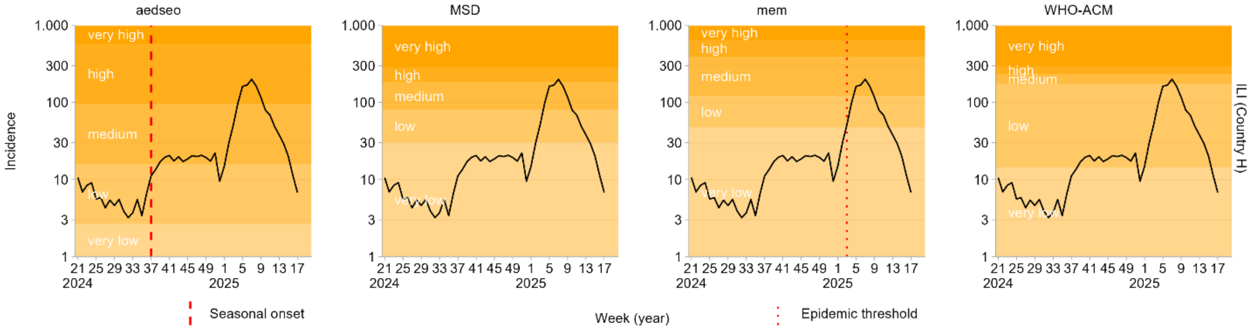
aedseo
MSD
mem
WHO-ACM
Incidence
ILI (Country H)
Seasonal onset
Week (year)
Epidemic threshold

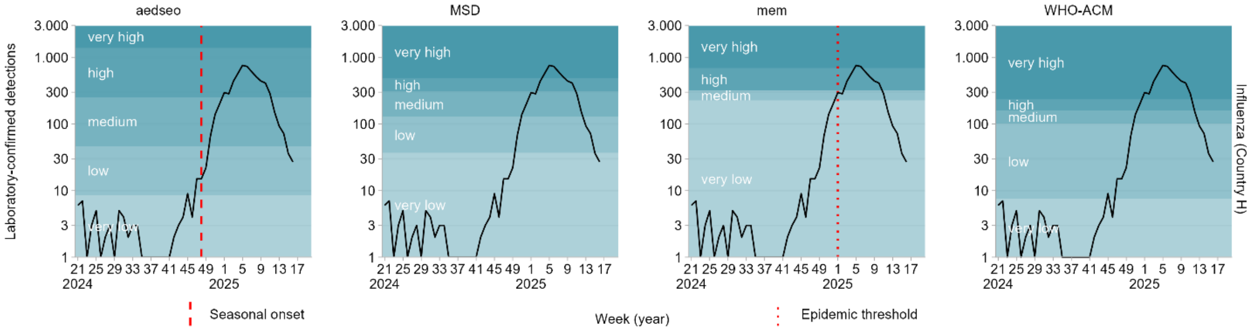
aedseo
MSD
mem
WHO-ACM
Laboratory-confirmed detections
Influenza (Country H)
Seasonal onset
Week (year)
Epidemic threshold

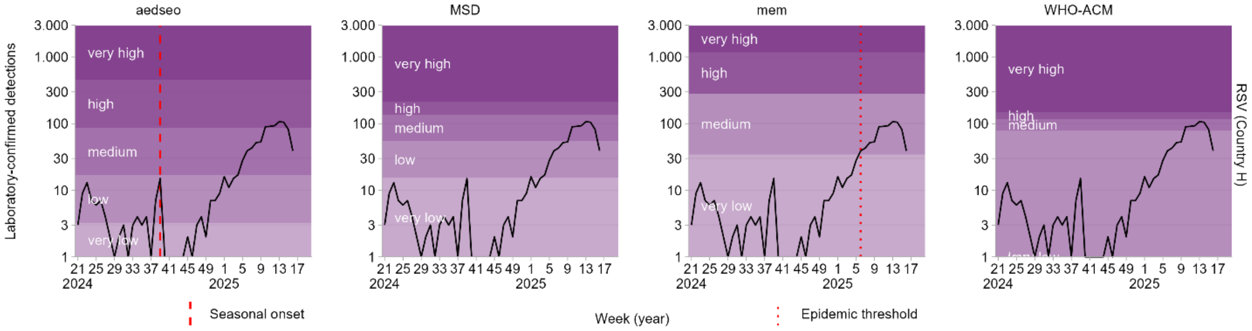
aedseo
MSD
mem
WHO-ACM
Laboratory-confirmed detections
RSV (Country H)
Seasonal onset
Week (year)
Epidemic threshold

### Country I

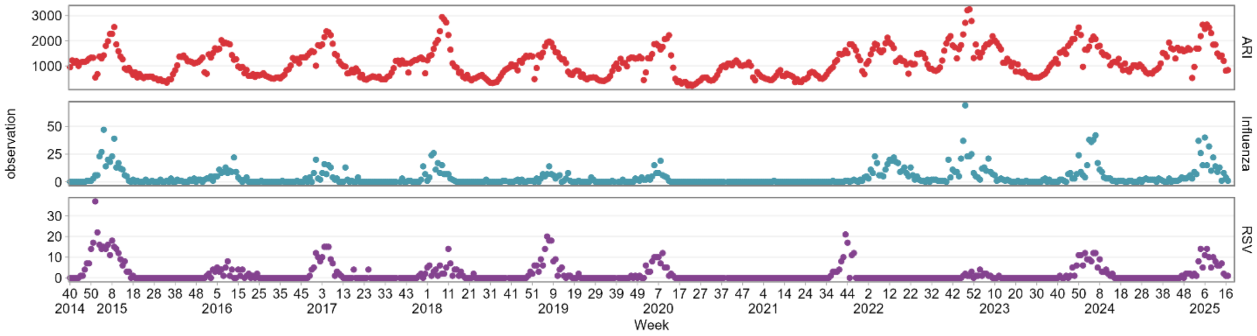
observation
ARI
Influenza
RSV
Week
2014 2015 2016 2017 2018 2019 2020 2021 2022 2023 2024 2025

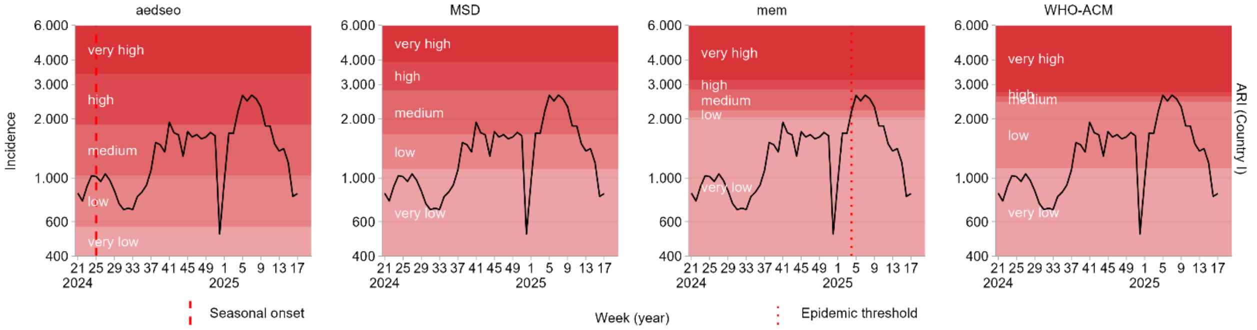
aedseo
MSD
mem
WHO-ACM
Incidence
very high
high
medium
low
very low
ARI (Country I)
Week (year)
Seasonal onset
Epidemic threshold

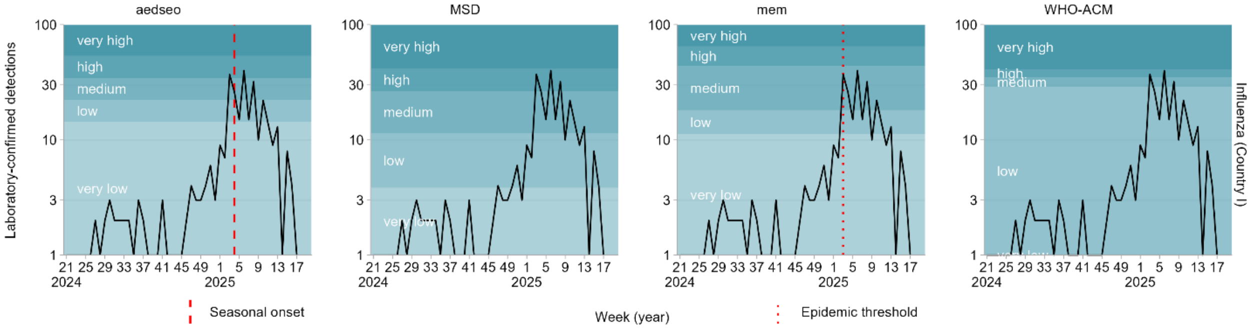
aedseo
MSD
mem
WHO-ACM
Laboratory-confirmed detections
very high
high
medium
low
very low
Influenza (Country I)
Week (year)
Seasonal onset
Epidemic threshold

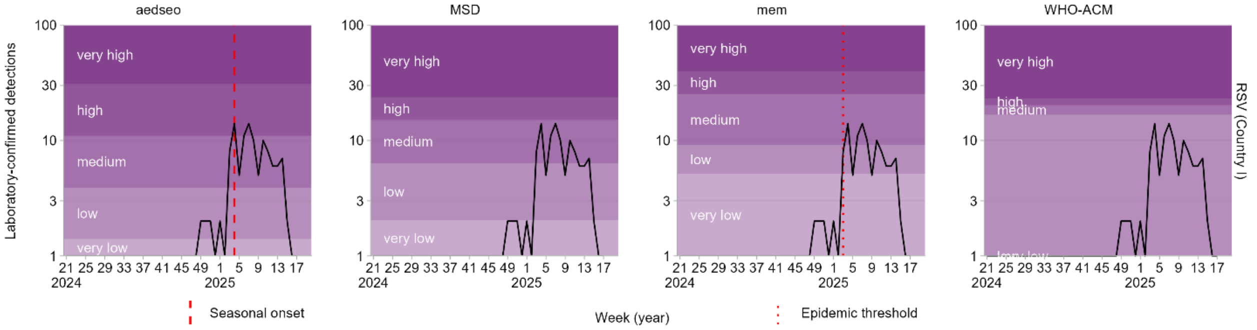
aedseo
MSD
mem
WHO-ACM
Laboratory-confirmed detections
very high
high
medium
low
very low
RSV (Country I)
Week (year)
Seasonal onset
Epidemic threshold

### Country J

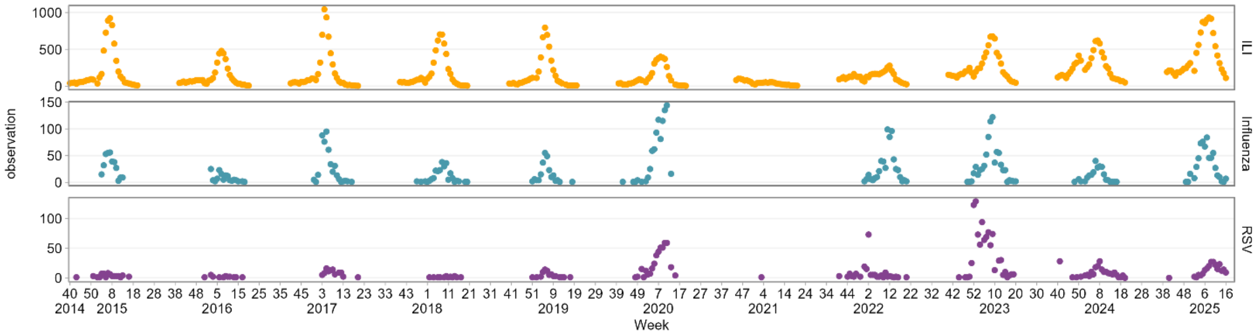

observation
ILI
Influenza
RSV
Week


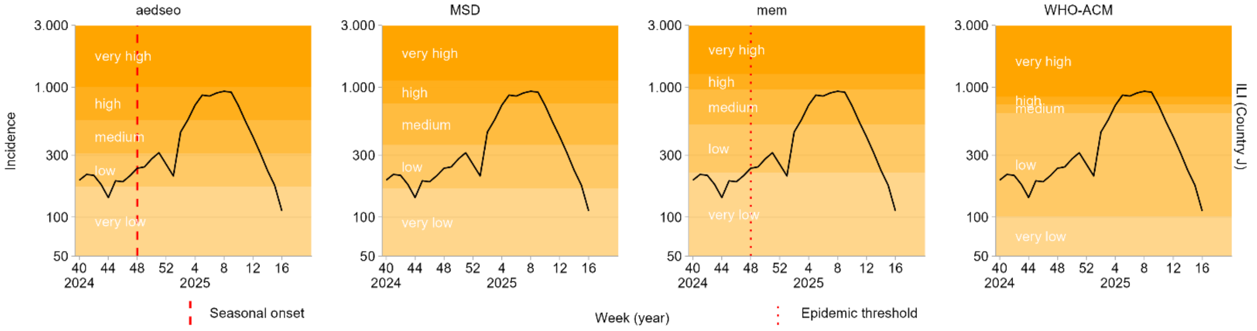

aedseo
MSD
mem
WHO-ACM
Incidence
ILI (Country J)
very high
high
medium
low
very low
Seasonal onset
Week (year)
Epidemic threshold


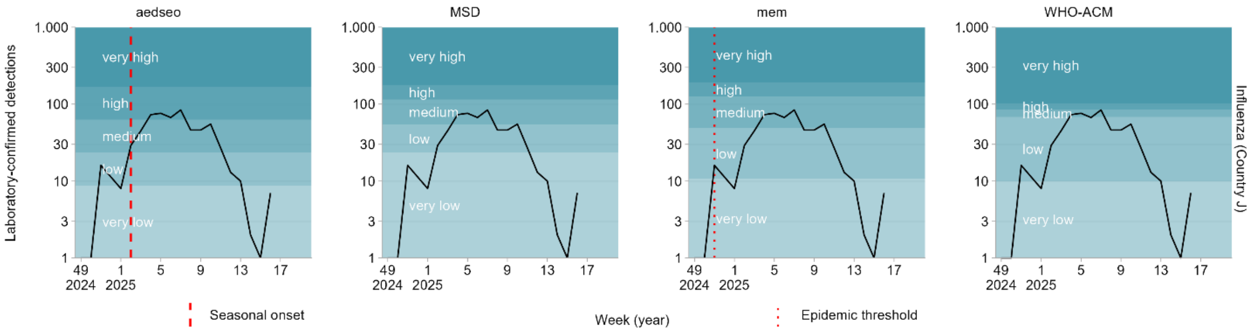

aedseo
MSD
mem
WHO-ACM
Laboratory-confirmed detections
Influenza (Country J)
very high
high
medium
low
very low
Seasonal onset
Week (year)
Epidemic threshold


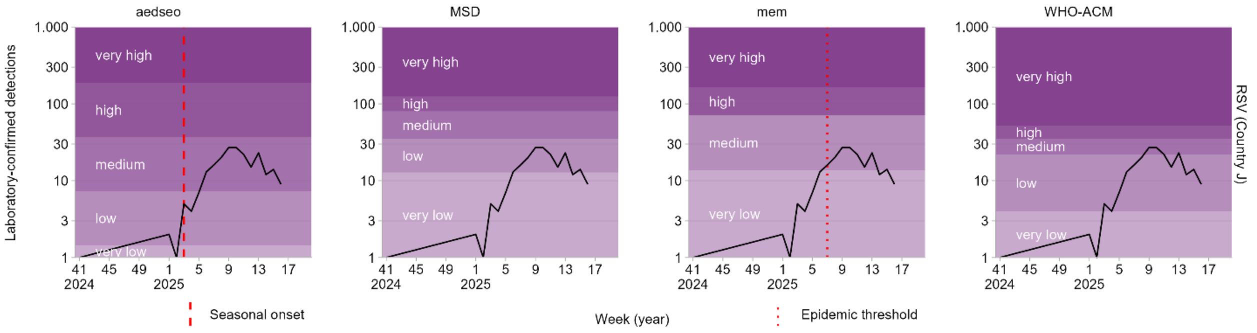

aedseo
MSD
mem
WHO-ACM
Laboratory-confirmed detections
RSV (Country J)
very high
high
medium
low
very low
Seasonal onset
Week (year)
Epidemic threshold

### Country K

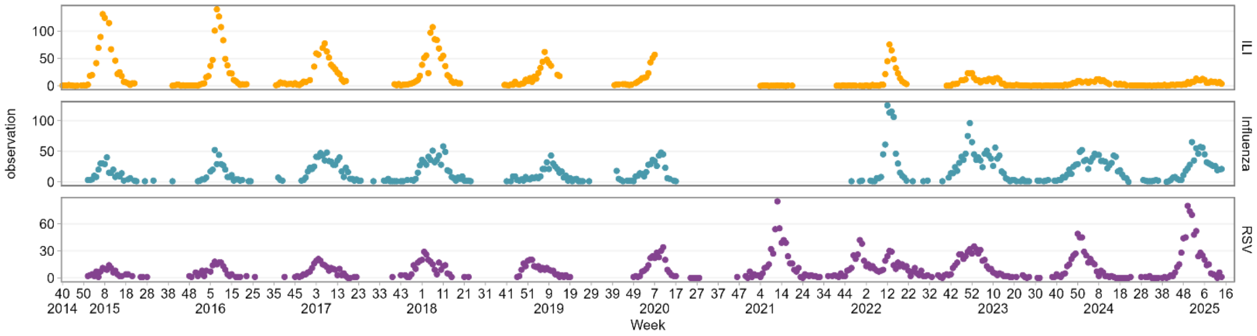

observation
ILI
Influenza
RSV
Week
2014 2015 2016 2017 2018 2019 2020 2021 2022 2023 2024 2025


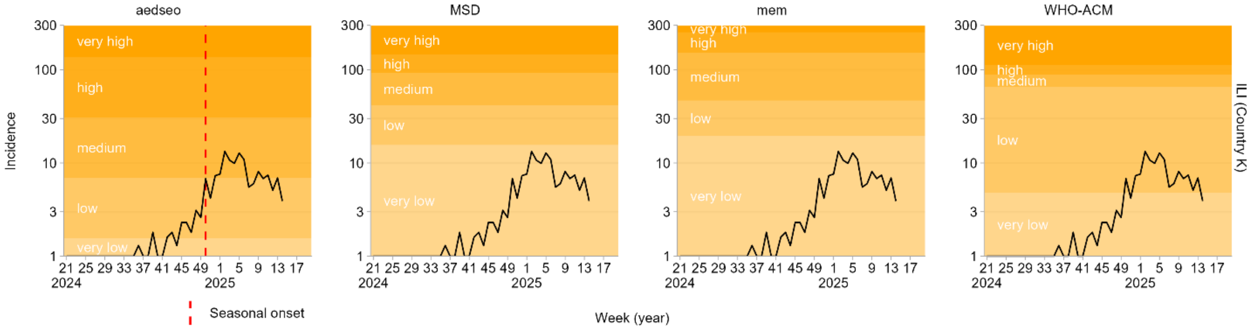

aedseo
MSD
mem
WHO-ACM
Incidence
ILI (Country K)
very high
high
medium
low
very low
Seasonal onset
Week (year)


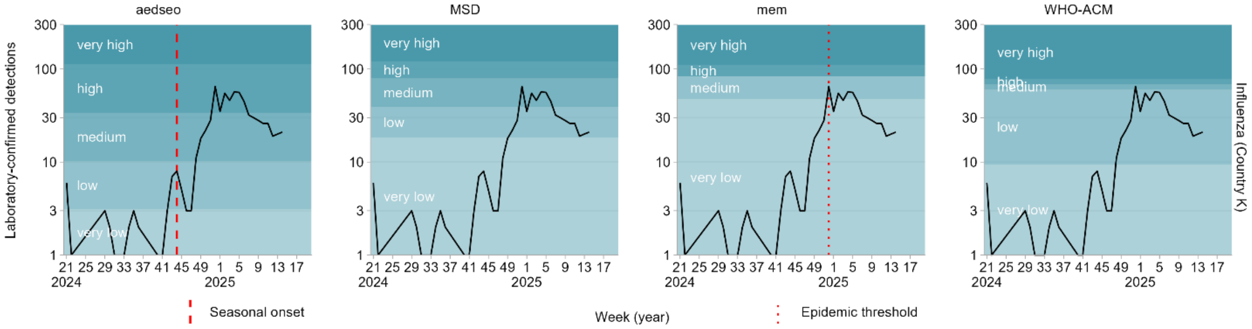

aedseo
MSD
mem
WHO-ACM
Laboratory-confirmed detections
Influenza (Country K)
very high
high
medium
low
very low
Seasonal onset
Week (year)
Epidemic threshold


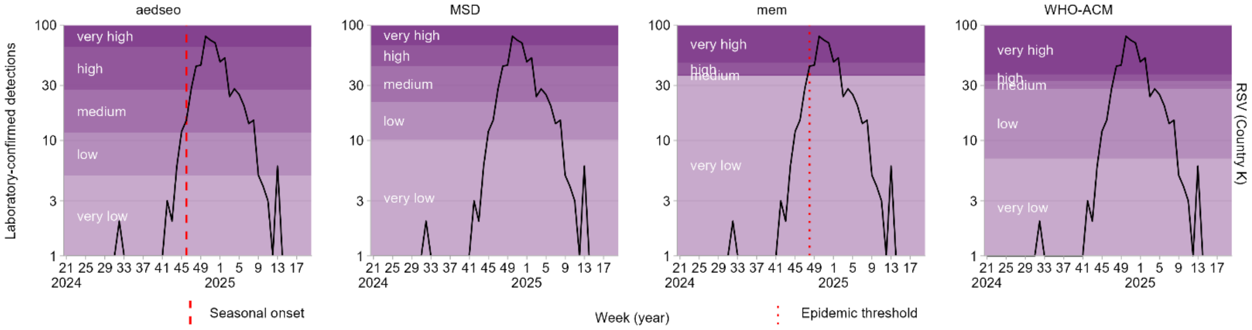

aedseo
MSD
mem
WHO-ACM
Laboratory-confirmed detections
RSV (Country K)
very high
high
medium
low
very low
Seasonal onset
Week (year)
Epidemic threshold

#### Country L

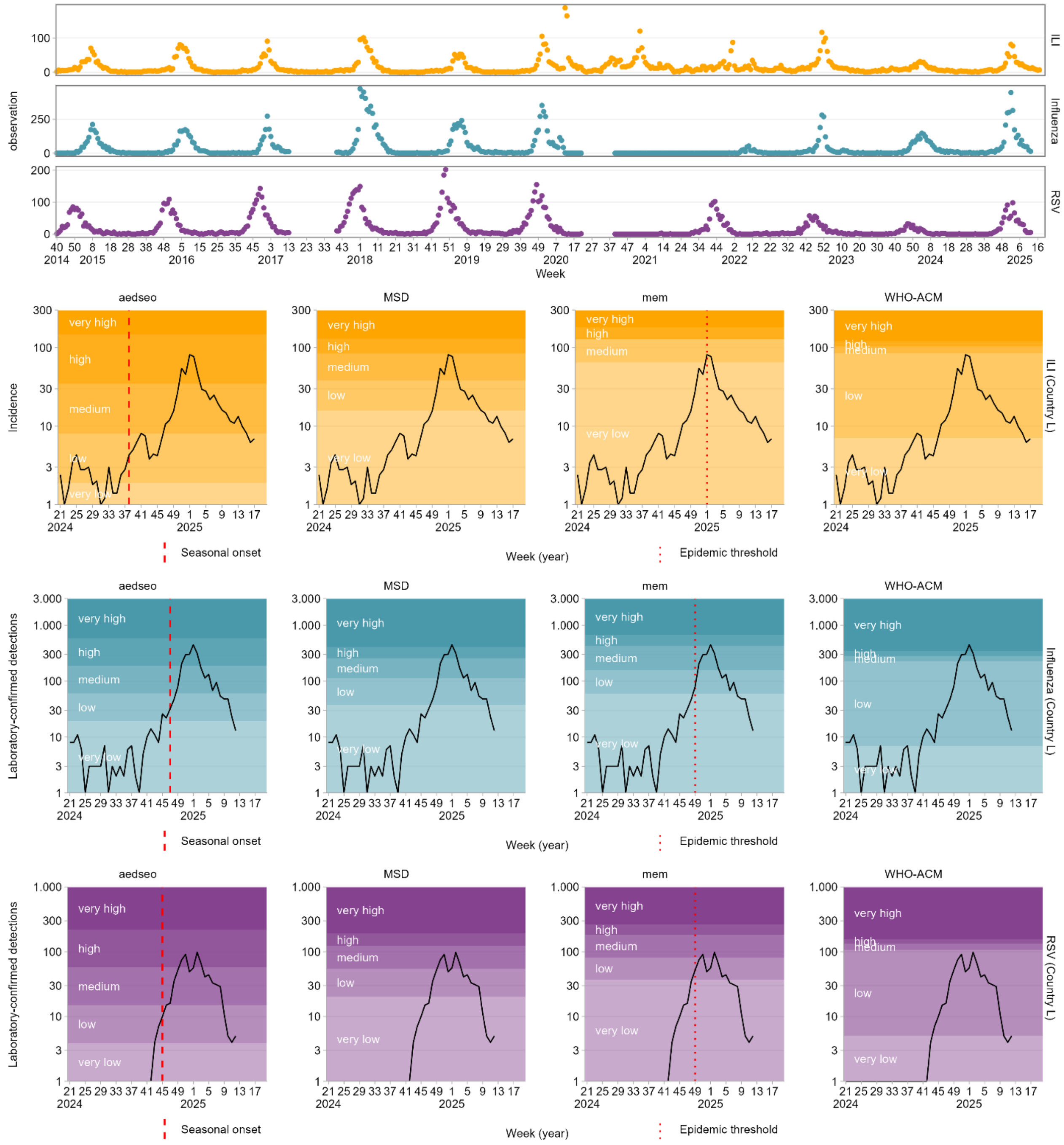

ILI
Influenza
RSV
observation
Week
aedseo
MSD
mem
WHO-ACM
Incidence
ILI (Country L)
very high
high
medium
low
very low
Seasonal onset
Week (year)
Epidemic threshold
Laboratory-confirmed detections
Influenza (Country L)
RSV (Country L)

#### Country M

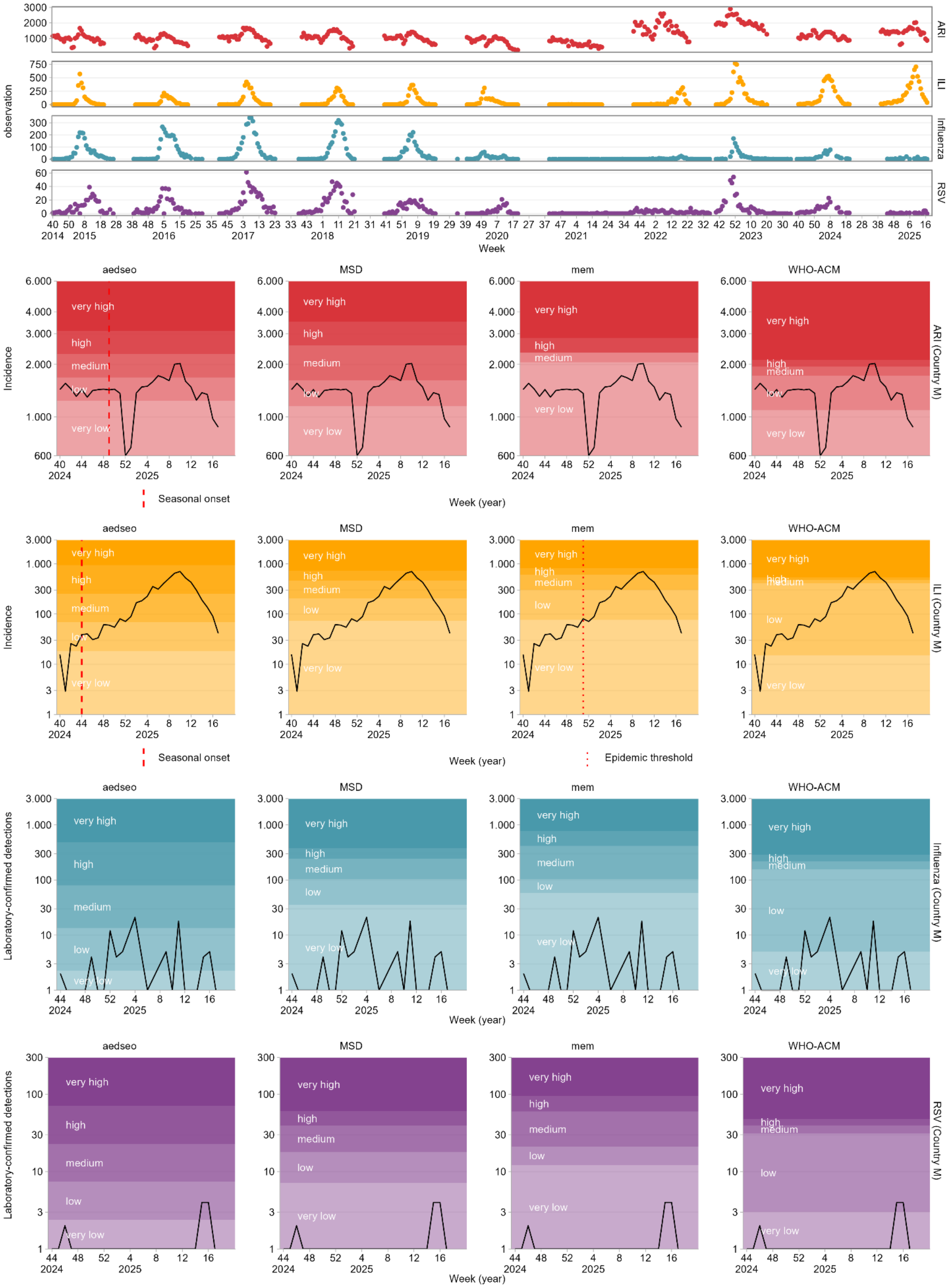

ARI
ILI
Influenza
RSV
observation
Week
aedseo
MSD
mem
WHO-ACM
very high
high
medium
low
very low
Incidence
Laboratory-confirmed detections
Seasonal onset
Epidemic threshold
Week (year)
ARI (Country M)
ILI (Country M)
Influenza (Country M)
RSV (Country M)

#### Country N

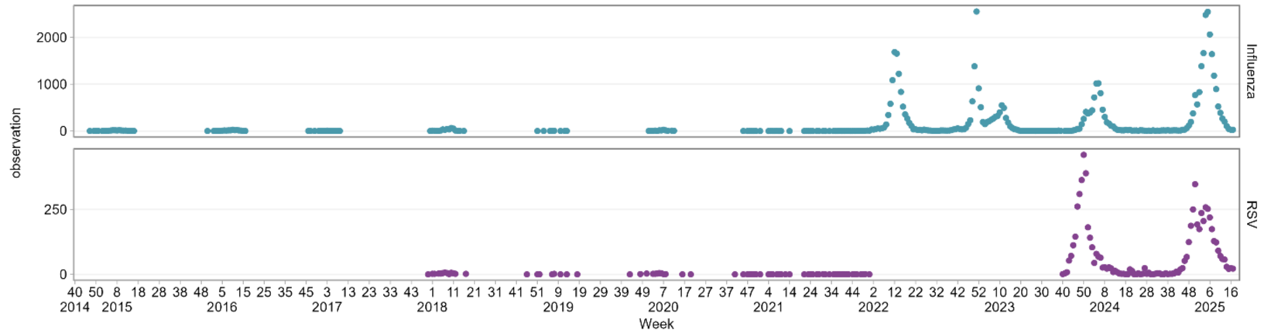
observation
Influenza
RSV
Week

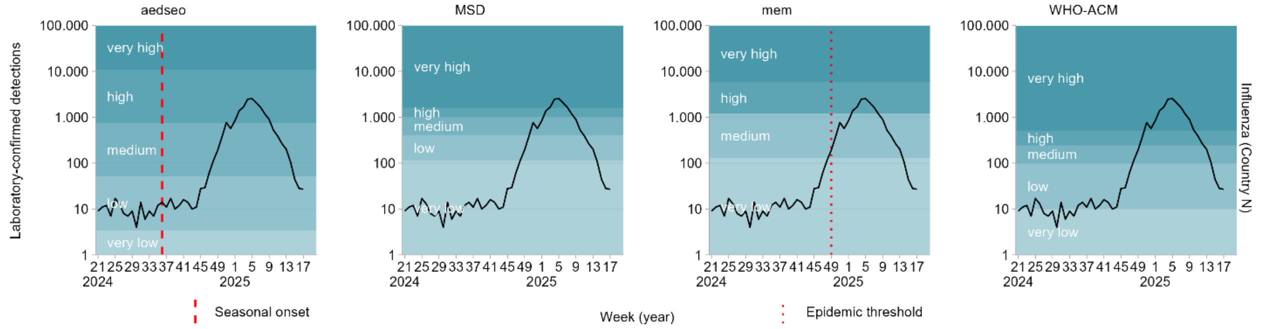
aedseo
MSD
mem
WHO-ACM
Laboratory-confirmed detections
Influenza (Country N)
very high
high
medium
low
very low
Week (year)
Seasonal onset
Epidemic threshold

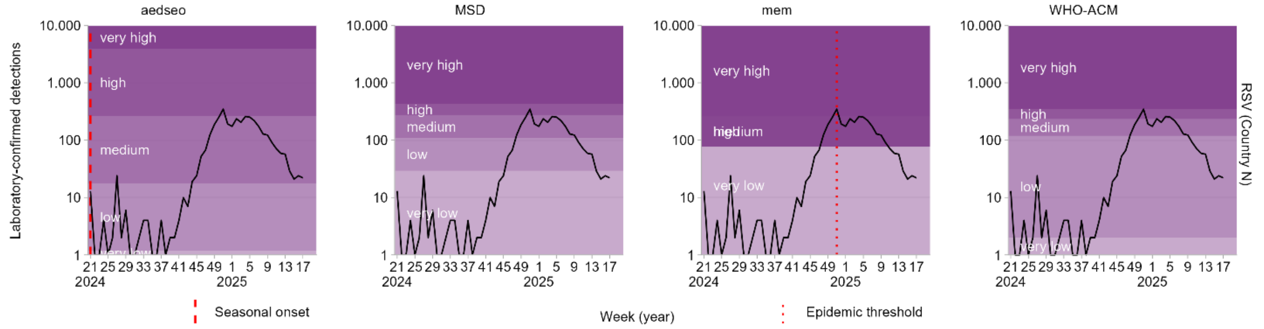
aedseo
MSD
mem
WHO-ACM
Laboratory-confirmed detections
RSV (Country N)
very high
high
medium
low
very low
Week (year)
Seasonal onset
Epidemic threshold

### Country O

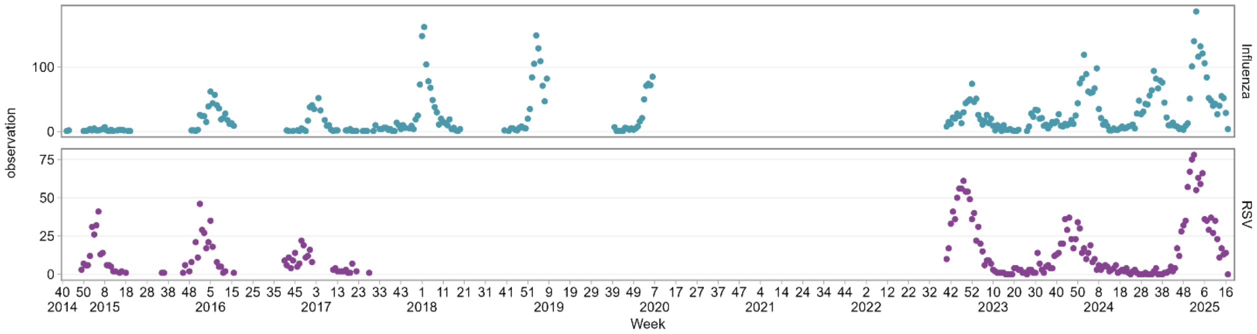
Influenza
RSV
observation
Week

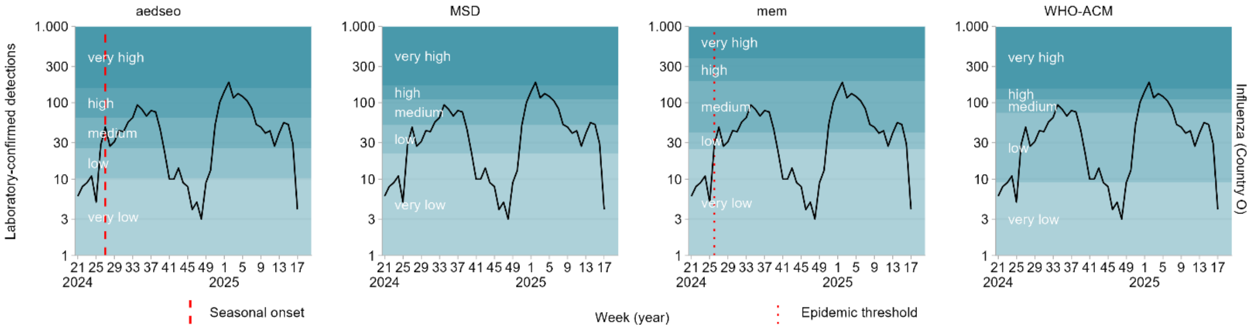
aedseo
MSD
mem
WHO-ACM
Laboratory-confirmed detections
Influenza (Country O)
very high
high
medium
low
very low
Seasonal onset
Week (year)
Epidemic threshold

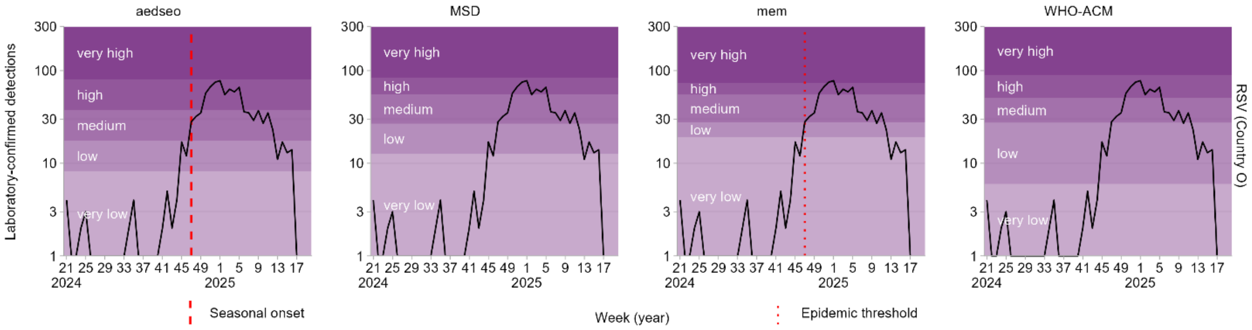
aedseo
MSD
mem
WHO-ACM
Laboratory-confirmed detections
RSV (Country O)
very high
high
medium
low
very low
Seasonal onset
Week (year)
Epidemic threshold

#### Country P

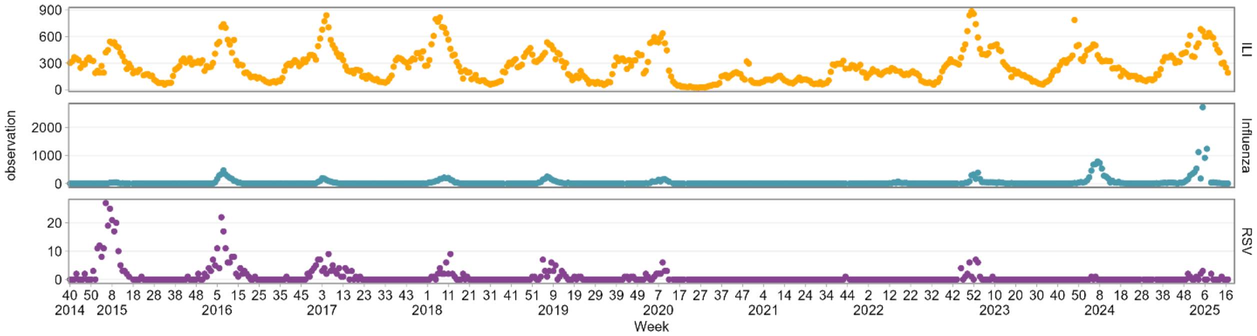

observation
ILI
Influenza
RSV
Week


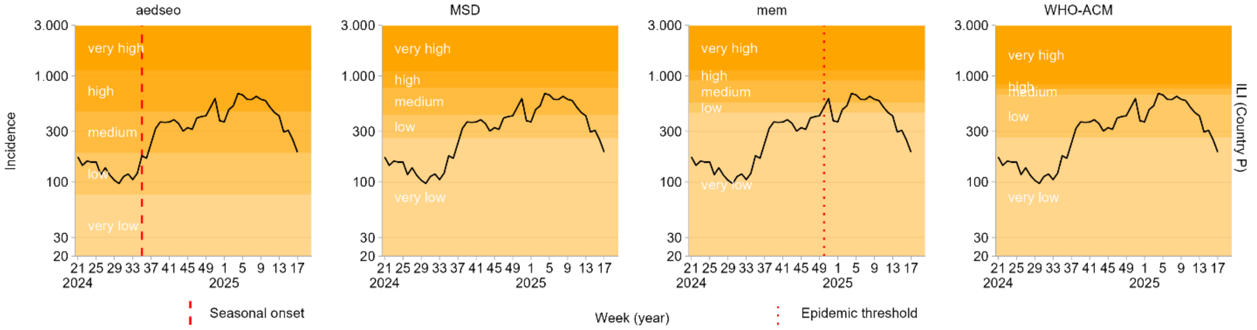

aedseo
MSD
mem
WHO-ACM
Incidence
ILI (Country P)
very high
high
medium
low
very low
Week (year)
Seasonal onset
Epidemic threshold


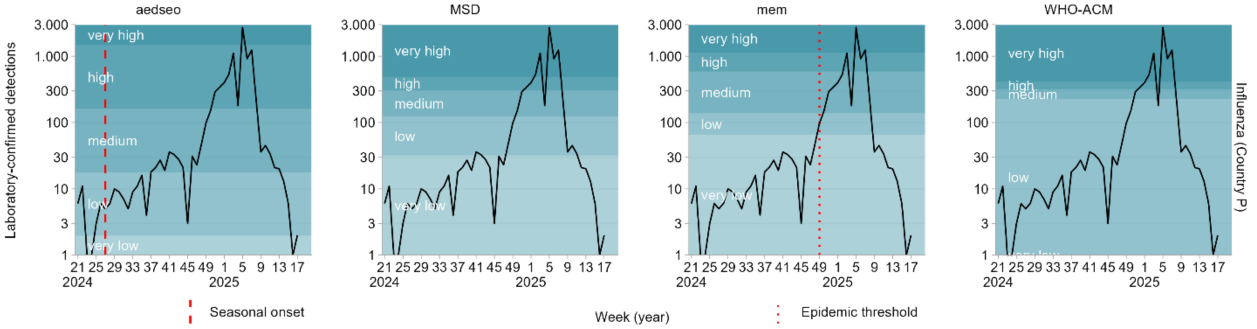

aedseo
MSD
mem
WHO-ACM
Laboratory-confirmed detections
Influenza (Country P)
very high
high
medium
low
very low
Week (year)
Seasonal onset
Epidemic threshold


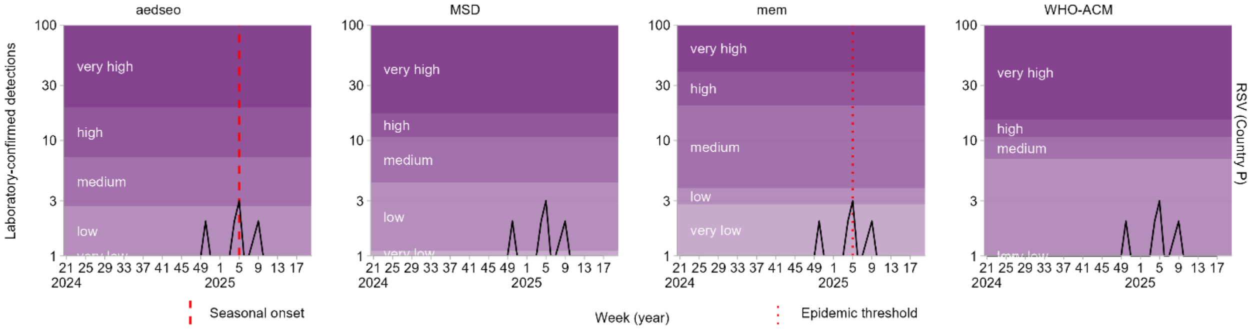

aedseo
MSD
mem
WHO-ACM
Laboratory-confirmed detections
RSV (Country P)
very high
high
medium
low
very low
Week (year)
Seasonal onset
Epidemic threshold

#### Country Q

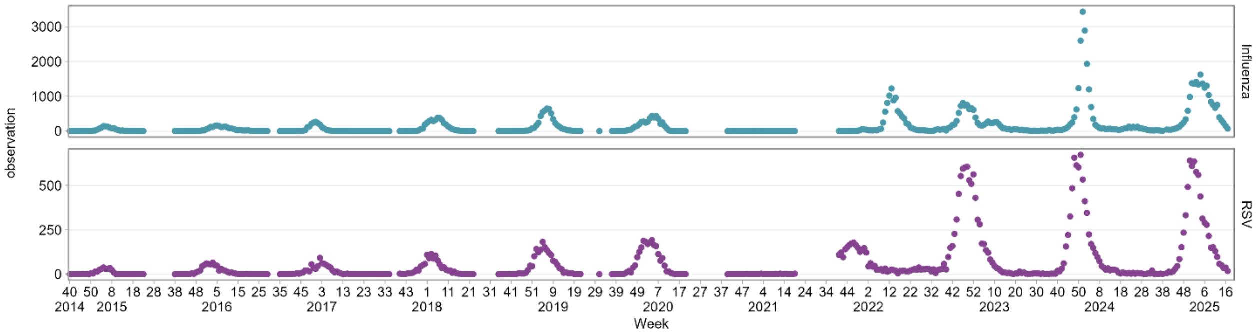
observation
3000
2000
1000
0
500
250
Influenza
RSV
40 50 8 18 28 38 48 5 15 25 35 45 3 13 23 33 43 1 11 21 31 41 51 9 19 29 39 49 7 17 27 37 47 4 14 24 34 44 2 12 22 32 42 52 10 20 30 40 50 8 18 28 38 48 6 16
2014 2015 2016 2017 2018 2019 2020 2021 2022 2023 2024 2025
Week

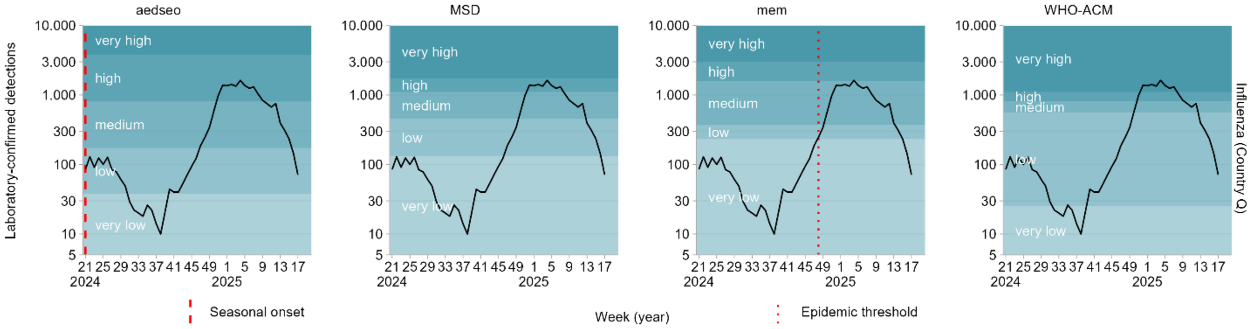
aedseo
MSD
mem
WHO-ACM
Laboratory-confirmed detections
10.000
3.000
1.000
300
100
30
10
5
very high
high
medium
low
very low
21 25 29 33 37 41 45 49 1 5 9 13 17
2024 2025
Influenza (Country Q)
Seasonal onset
Week (year)
Epidemic threshold

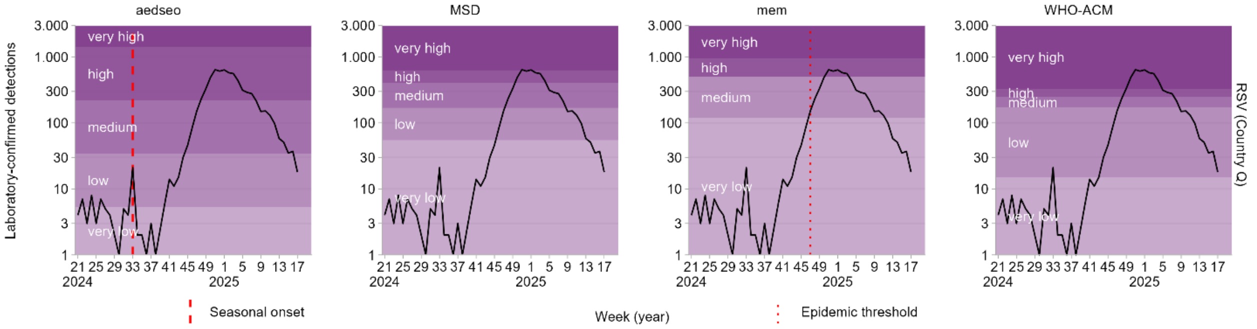
aedseo
MSD
mem
WHO-ACM
Laboratory-confirmed detections
3.000
1.000
300
100
30
10
3
1
very high
high
medium
low
very low
21 25 29 33 37 41 45 49 1 5 9 13 17
2024 2025
RSV (Country Q)
Seasonal onset
Week (year)
Epidemic threshold

#### Country R

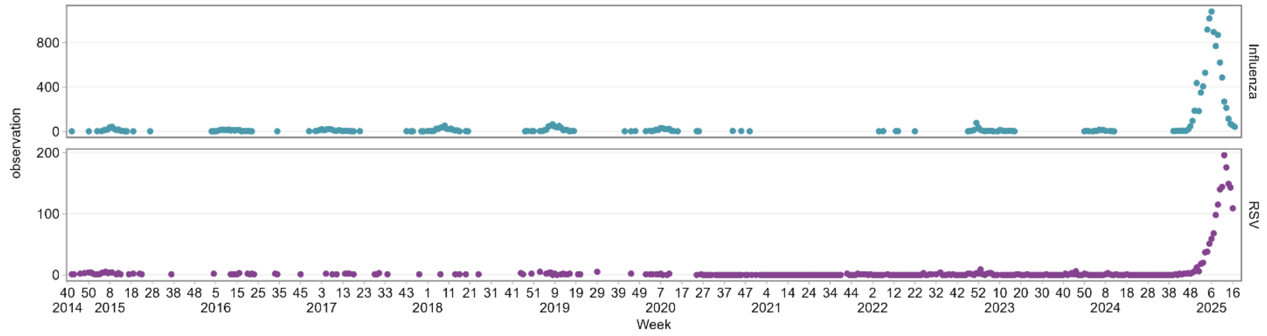

observation
800
400
0
200
100
Influenza
RSV
40 50 8 18 28 38 48 5 15 25 35 45 3 13 23 33 43 1 11 21 31 41 51 9 19 29 39 49 7 17 27 37 47 4 14 24 34 44 2 12 22 32 42 52 10 20 30 40 50 8 18 28 38 48 6 16
2014 2015 2016 2017 2018 2019 2020 2021 2022 2023 2024 2025
Week


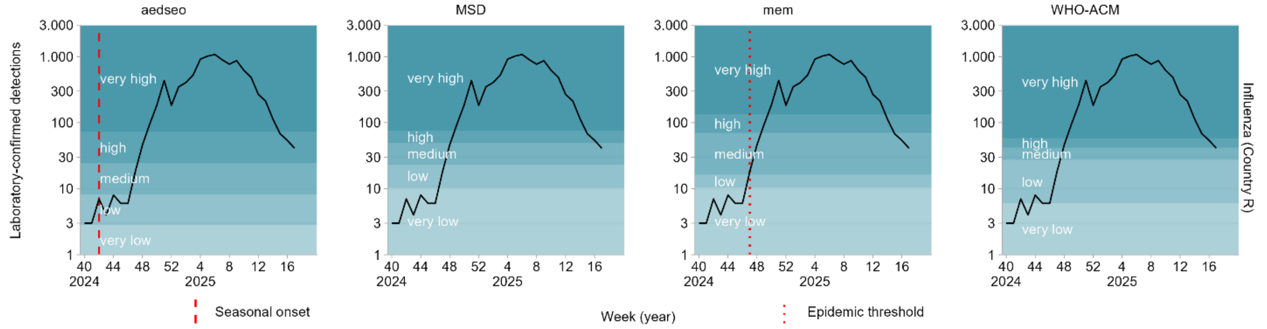

aedseo
MSD
mem
WHO-ACM
Laboratory-confirmed detections
3.000
1.000
300
100
30
10
3
1
very high
high
medium
low
very low
40 44 48 52 4 8 12 16
2024 2025
Influenza (Country R)
Seasonal onset
Week (year)
Epidemic threshold


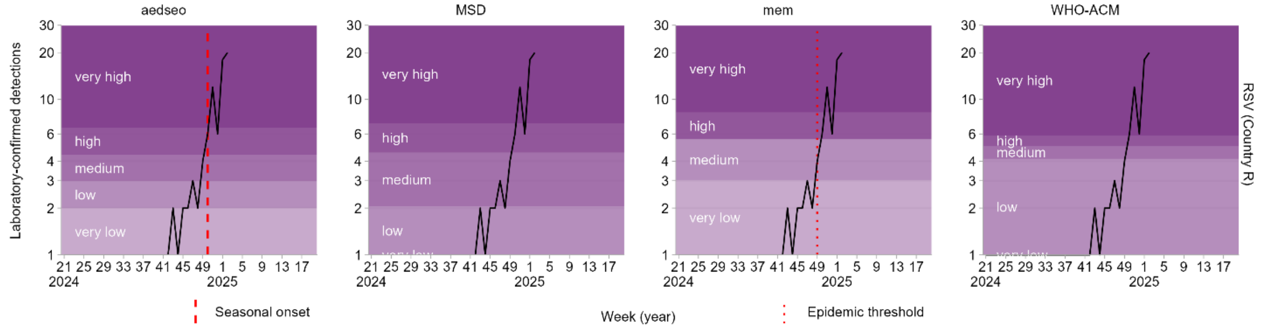

aedseo
MSD
mem
WHO-ACM
Laboratory-confirmed detections
30
20
10
6
4
3
2
1
very high
high
medium
low
very low
21 25 29 33 37 41 45 49 1 5 9 13 17
2024 2025
RSV (Country R)
Seasonal onset
Week (year)
Epidemic threshold

### Country S

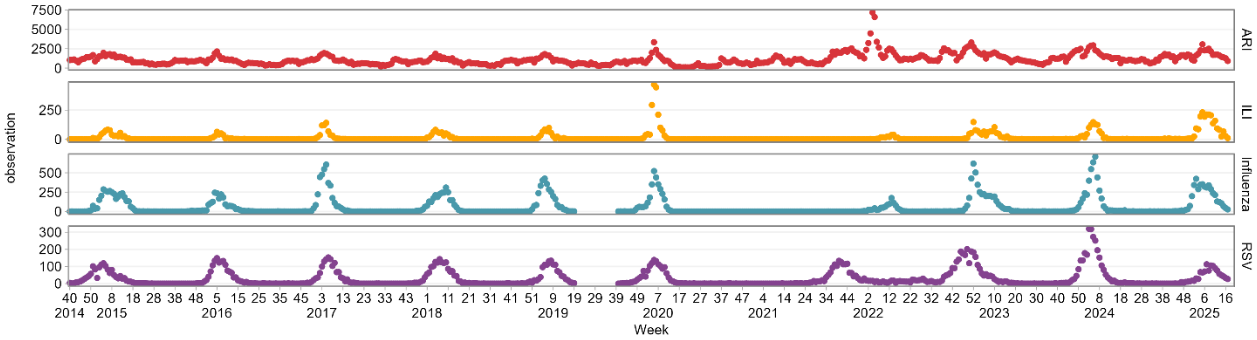
ARI
ILI
Influenza
RSV
observation
Week
7500
5000
2500
0
250
500
300
200
100
40 50 8 18 28 38 48 5 15 25 35 45 3 13 23 33 43 1 11 21 31 41 51 9 19 29 39 49 7 17 27 37 47 4 14 24 34 44 2 12 22 32 42 52 10 20 30 40 50 8 18 28 38 48 6 16
2014 2015 2016 2017 2018 2019 2020 2021 2022 2023 2024 2025

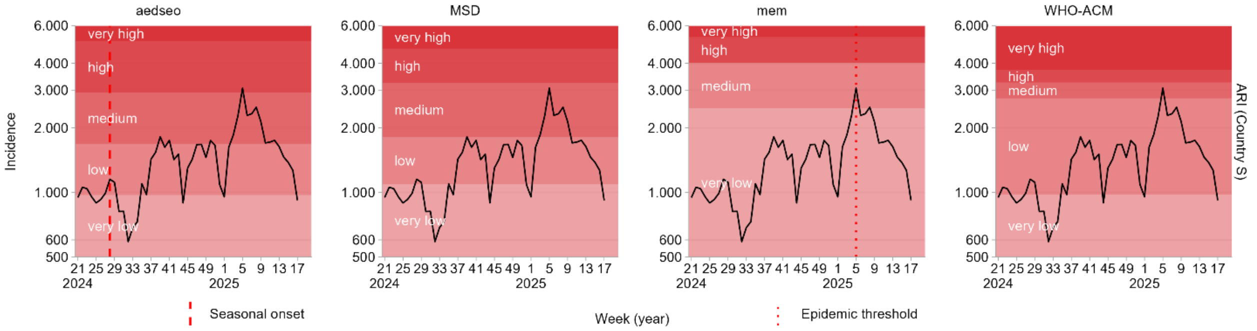
aedseo
MSD
mem
WHO-ACM
ARI (Country S)
Incidence
very high
high
medium
low
very low
6.000
4.000
3.000
2.000
1.000
600
500
21 25 29 33 37 41 45 49 1 5 9 13 17
2024
2025
Seasonal onset
Week (year)
Epidemic threshold

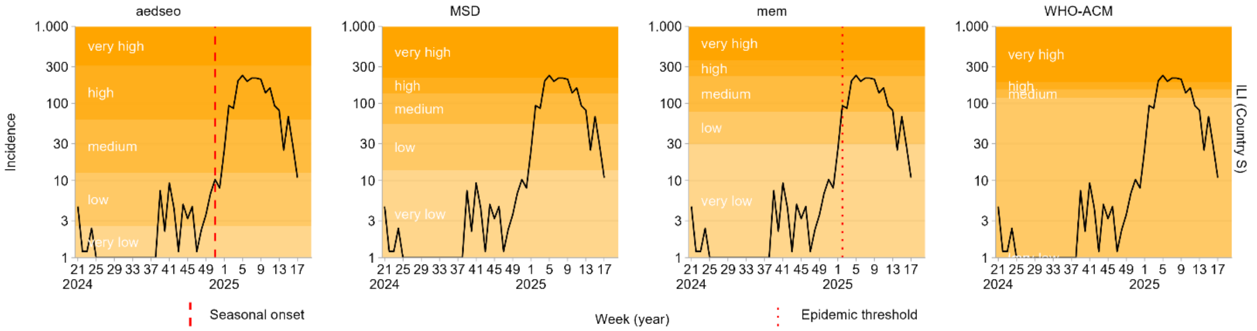
aedseo
MSD
mem
WHO-ACM
ILI (Country S)
Incidence
very high
high
medium
low
very low
1.000
300
100
30
10
3
1
21 25 29 33 37 41 45 49 1 5 9 13 17
2024
2025
Seasonal onset
Week (year)
Epidemic threshold

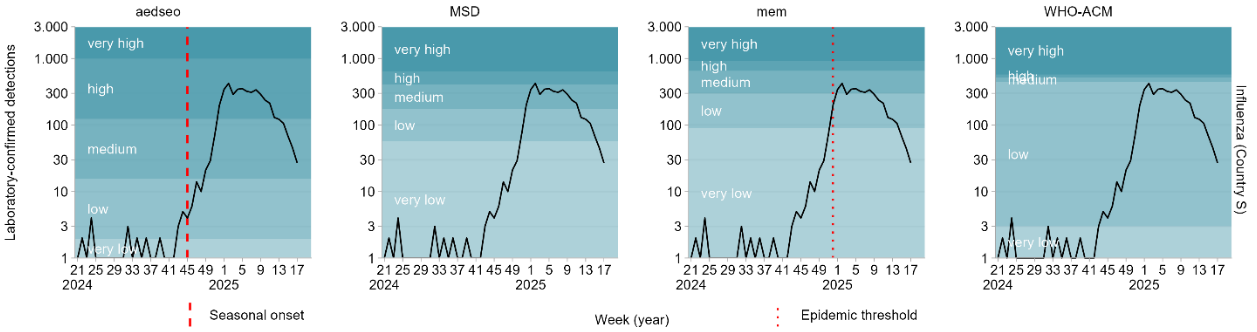
aedseo
MSD
mem
WHO-ACM
Influenza (Country S)
Laboratory-confirmed detections
very high
high
medium
low
very low
3.000
1.000
300
100
30
10
3
1
21 25 29 33 37 41 45 49 1 5 9 13 17
2024
2025
Seasonal onset
Week (year)
Epidemic threshold

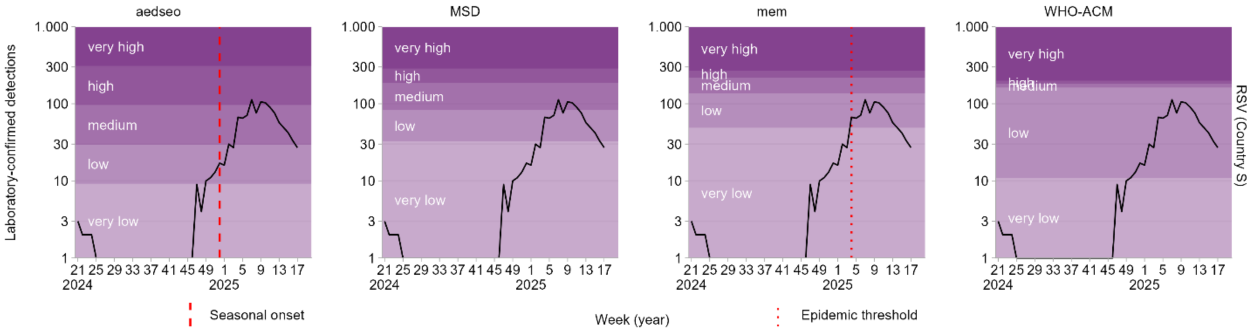
aedseo
MSD
mem
WHO-ACM
RSV (Country S)
Laboratory-confirmed detections
very high
high
medium
low
very low
1.000
300
100
30
10
3
1
21 25 29 33 37 41 45 49 1 5 9 13 17
2024
2025
Seasonal onset
Week (year)
Epidemic threshold

#### Country T

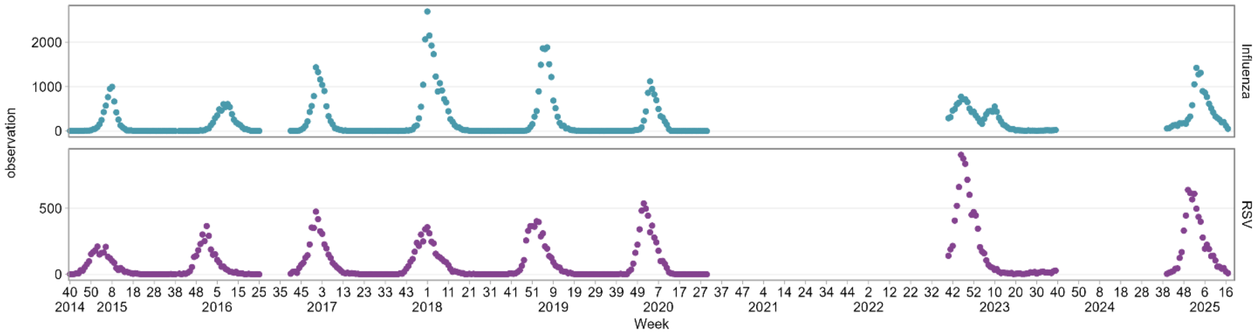

observation
Influenza
RSV
2000
1000
0
500
0
40 50 8 18 28 38 48 5 15 25 35 45 3 13 23 33 43 1 11 21 31 41 51 9 19 29 39 49 7 17 27 37 47 4 14 24 34 44 2 12 22 32 42 52 10 20 30 40 50 8 18 28 38 48 6 16
2014 2015 2016 2017 2018 2019 2020 2021 2022 2023 2024 2025
Week


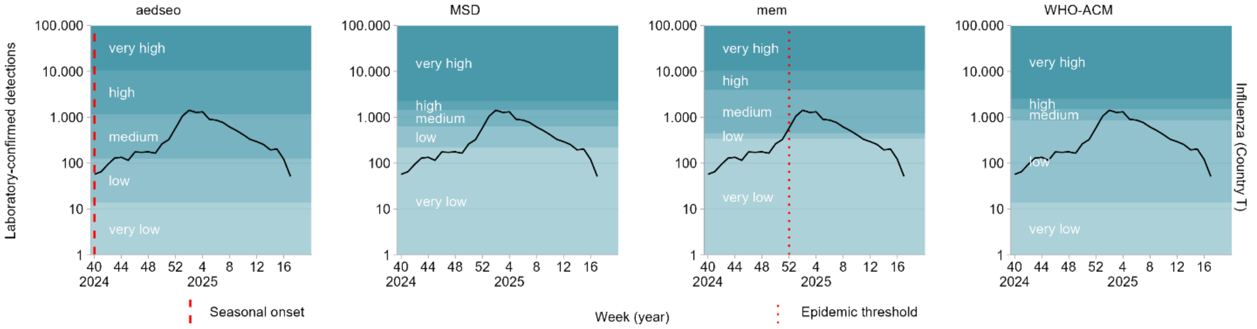

aedseo
MSD
mem
WHO-ACM
Laboratory-confirmed detections
Influenza (Country T)
100.000
10.000
1.000
100
10
1
very high
high
medium
low
very low
40 44 48 52 4 8 12 16
2024 2025
Seasonal onset
Week (year)
Epidemic threshold


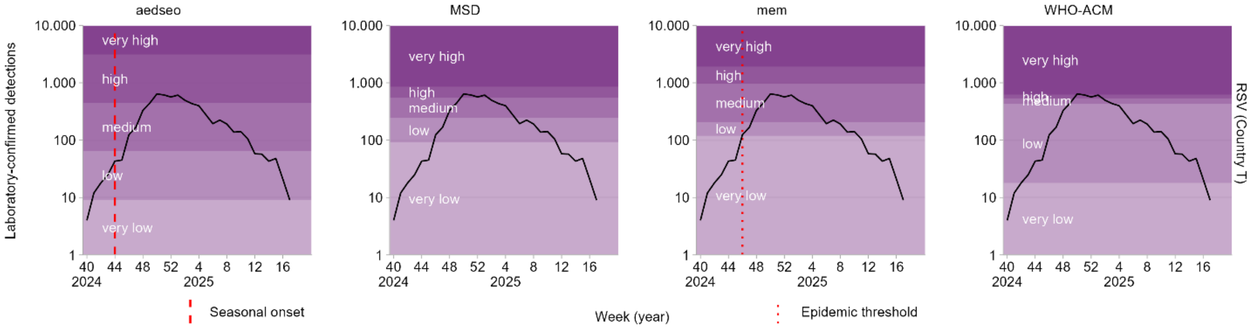

aedseo
MSD
mem
WHO-ACM
Laboratory-confirmed detections
RSV (Country T)
10.000
1.000
100
10
1
very high
high
medium
low
very low
40 44 48 52 4 8 12 16
2024 2025
Seasonal onset
Week (year)
Epidemic threshold

### Country U

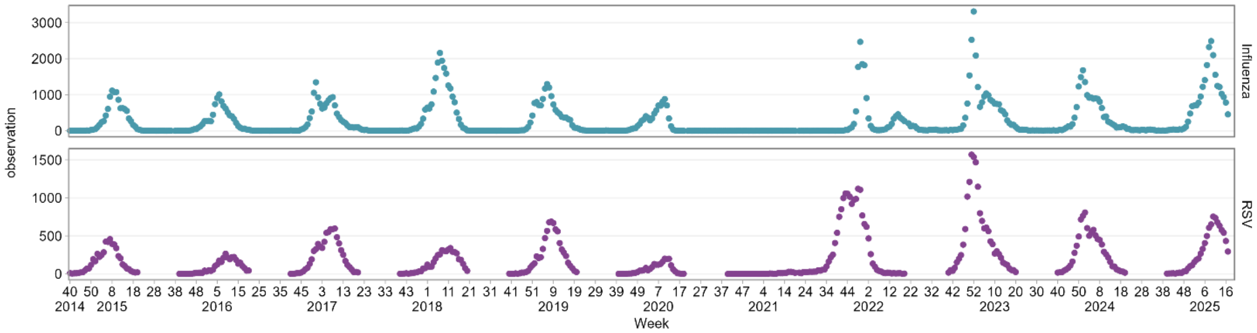
observation
Influenza
RSV
Week
2014
2015
2016
2017
2018
2019
2020
2021
2022
2023
2024
2025

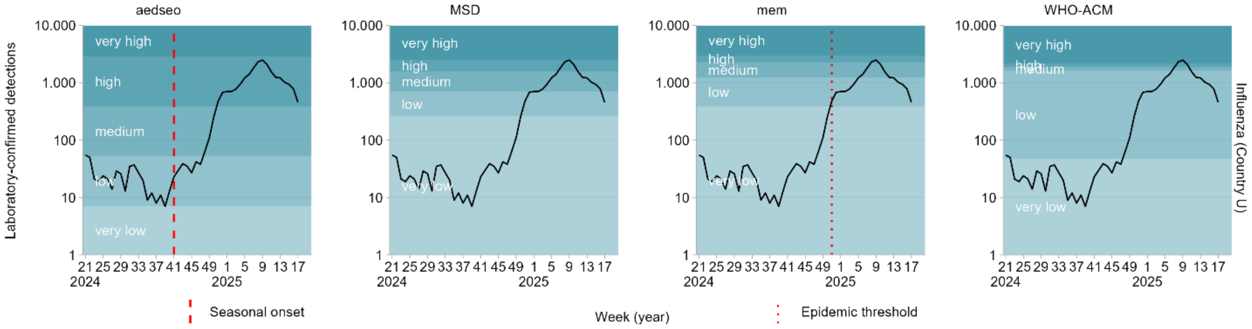
aedseo
MSD
mem
WHO-ACM
Laboratory-confirmed detections
very high
high
medium
low
very low
Influenza (Country U)
Week (year)
Seasonal onset
Epidemic threshold

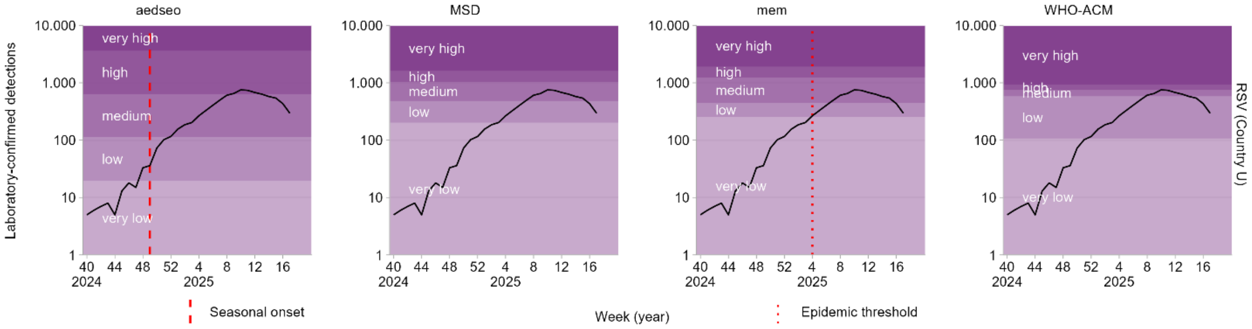
aedseo
MSD
mem
WHO-ACM
Laboratory-confirmed detections
very high
high
medium
low
very low
RSV (Country U)
Week (year)
Seasonal onset
Epidemic threshold

#### Supplement S2

| Country | Disease | aedseo disease-specific threshold | mem epidemic threshold |
|---|---|---|---|
| A | Influenza | 42.40 | 356.47 |
| A | RSV | 4.87 | 501.47 |
| A | ILI | 40.46 | 154.28 |
| B | Influenza | 53.51 | 272.76 |
| B | RSV | 10.84 | 110.53 |
| B | ILI | 14.93 | 73.17 |
| C | Influenza | 11.84 | 758.45 |
| C | RSV | 19.28 | 424.06 |
| C | ARI | 62.20 | 242.14 |
| C | ILI | 22.75 | 124.09 |
| D | Influenza | 2.25 | 41.97 |
| D | RSV | 2.20 | 11.39 |
| D | ARI | 75.84 | 427.67 |
| D | ILI | 126.07 | 259.81 |
| E | Influenza | 5.47 | 335.84 |
| E | RSV | 25.86 | 542.54 |
| E | ARI | 556.31 | 2194.27 |
| E | ILI | 122.62 | 398.34 |
| F | Influenza | 12.40 | 30.41 |
| F | RSV | 2.80 | 16.84 |
| F | ARI | 240.64 | 1255.68 |
| G | Influenza | 6.79 | 102.15 |
| G | RSV | 13.26 | 9.58 |
| G | ILI | 1.00 | 23.38 |
| H | Influenza | 8.55 | 224.93 |
| H | RSV | 3.25 | 34.59 |
| H | ARI | 291.98 | 1281.14 |
| H | ILI | 2.66 | 46.90 |
| I | Influenza | 14.40 | 11.25 |
| I | RSV | 1.40 | 5.12 |
| I | ARI | 565.63 | 2031.16 |
| J | Influenza | 8.74 | 10.65 |
| J | RSV | 1.45 | 13.76 |
| J | ILI | 170.79 | 219.97 |

| | | | |
|---|---|---|---|
| K | Influenza | 3.11 | 47.77 |
| K | RSV | 4.96 | 37.32 |
| K | ILI | 1.56 | 19.38 |
| L | Influenza | 19.45 | 59.46 |
| L | RSV | 3.94 | 37.07 |
| L | ILI | 1.88 | 64.77 |
| M | Influenza | 2.23 | 57.75 |
| M | RSV | 2.39 | 12.15 |
| M | ARI | 1240.99 | 2059.95 |
| M | ILI | 18.47 | 76.44 |
| N | Influenza | 3.46 | 129.00 |
| N | RSV | 1.20 | 266.68 |
| O | Influenza | 10.31 | 24.49 |
| O | RSV | 8.22 | 19.11 |
| P | Influenza | 1.97 | 65.19 |
| P | RSV | 1.00 | 2.80 |
| P | ILI | 76.53 | 450.43 |
| Q | Influenza | 37.44 | 231.98 |
| Q | RSV | 5.33 | 120.34 |
| R | Influenza | 2.80 | 10.40 |
| R | RSV | 2.00 | 3.02 |
| S | Influenza | 1.94 | 89.57 |
| S | RSV | 9.23 | 49.11 |
| S | ARI | 971.02 | 2464.71 |
| S | ILI | 2.55 | 29.80 |
| T | Influenza | 13.71 | 339.28 |
| T | RSV | 9.20 | 118.99 |
| U | Influenza | 7.19 | 393.75 |
| U | RSV | 19.83 | 256.87 |

#### Supplement S3

| Country | Disease | Seasonal peak (week) | aedseo seasonal onset (week) | mem epi. threshold (week) | No. weeks between seasonal onset vs. mem epi. threshold | No. weeks with significant growth | No. weeks with no significant growth |
|---|---|---|---|---|---|---|---|
| A | Influenza | 9 | 48 | 51 | 3 | 3 | 0 |
| A | RSV | 6 | 46 | 4 | 10 | 10 | 0 |
| B | Influenza | 5 | 49 | 51 | 2 | 2 | 0 |
| B | RSV | 1 | 45 | 48 | 3 | 3 | 0 |
| B | ILI | 5 | 2 | 4 | 2 | 2 | 0 |
| C | Influenza | 1 | 44 | 50 | 6 | 6 | 0 |
| C | RSV | 51 | 41 | 46 | 5 | 5 | 0 |
| C | ARI | 4 | 36 | 38 | 2 | 2 | 0 |
| C | ILI | 4 | 36 | 38 | 2 | 2 | 0 |
| D | ILI | 5 | 37 | 41 | 4 | 4 | 0 |
| E | Influenza | 4 | 25 | 49 | 24 | 10 | 14 |
| E | RSV | 50 | 26 | 47 | 21 | 11 | 10 |
| E | ILI | 4 | 41 | 1 | 12 | 6 | 6 |
| F | Influenza | 4 | 1 | 2 | 1 | 1 | 0 |
| F | RSV | 8 | 4 | 8 | 4 | 4 | 0 |
| F | ARI | 4 | 33 | 3 | 22 | 8 | 14 |
| G | Influenza | 5 | 51 | 2 | 3 | 3 | 0 |
| G | ILI | 5 | 51 | 2 | 3 | 3 | 0 |
| H | Influenza | 5 | 48 | 1 | 5 | 5 | 0 |
| H | RSV | 13 | 39 | 6 | 19 | 8 | 11 |
| H | ARI | 5 | 36 | 47 | 11 | 7 | 4 |
| H | ILI | 7 | 37 | 3 | 18 | 6 | 12 |
| I | ARI | 7 | 25 | 4 | 31 | 8 | 23 |
| J | RSV | 9 | 3 | 7 | 4 | 4 | 0 |

| | | | | | | | |
|---|---|---|---|---|---|---|---|
| K | Influenza | 52 | 44 | 52 | 8 | 4 | 4 |
| K | RSV | 50 | 46 | 48 | 2 | 2 | 0 |
| L | Influenza | 1 | 47 | 49 | 2 | 2 | 0 |
| L | RSV | 2 | 45 | 49 | 4 | 4 | 0 |
| L | ILI | 1 | 38 | 1 | 15 | 11 | 4 |
| M | ILI | 10 | 44 | 51 | 7 | 4 | 3 |
| N | Influenza | 5 | 36 | 49 | 13 | 5 | 8 |
| N | RSV | 51 | 21 | 51 | 30 | 12 | 18 |
| P | Influenza | 5 | 27 | 49 | 22 | 6 | 16 |
| P | ILI | 4 | 35 | 50 | 15 | 10 | 5 |
| Q | Influenza | 4 | 21 | 48 | 26 | 7 | 19 |
| Q | RSV | 51 | 33 | 47 | 13 | 7 | 6 |
| R | Influenza | 6 | 42 | 47 | 5 | 2 | 3 |
| S | Influenza | 2 | 45 | 52 | 7 | 7 | 0 |
| S | RSV | 7 | 52 | 4 | 4 | 4 | 0 |
| S | ARI | 5 | 28 | 5 | 29 | 9 | 20 |
| S | ILI | 5 | 51 | 2 | 3 | 3 | 0 |
| T | Influenza | 2 | 40 | 52 | 12 | 10 | 2 |
| T | RSV | 50 | 44 | 46 | 2 | 2 | 0 |
| U | Influenza | 9 | 41 | 51 | 10 | 7 | 3 |
| U | RSV | 10 | 49 | 4 | 7 | 7 | 0 |

Table S3-1. Shows 45 data sources with the time (weeks) from when aedseo estimates seasonal onset to when mem crosses its epidemic threshold, and how many of these weeks have significant growth or not.

| Country | Disease | Seasonal peak (week) | mem epi. threshold (week) | aedseo seasonal onset (week) | No. weeks between epi. threshold vs. seasonal onset | No. weeks with significant growth | No. weeks with no significant growth |
|---|---|---|---|---|---|---|---|
| A | ILI | 7 | 29 | 31 | 2 | 1 | 1 |
| G | RSV | 7 | 48 | 51 | 3 | 3 | 0 |
| I | Influenza | 6 | 3 | 4 | 1 | 1 | 0 |
| I | RSV | 4 | 3 | 4 | 1 | 1 | 0 |
| J | Influenza | 7 | 51 | 2 | 2 | 1 | 1 |
| O | Influenza | 2 | 26 | 27 | 1 | 1 | 0 |
| R | RSV | 12 | 49 | 50 | 1 | 1 | 0 |

Table S3-2. Shows seven data sources with the time (weeks) from when mem has crossed its epidemic threshold to when aedseo estimates seasonal onset, and how many of these weeks have significant growth or not.

| Country | Disease | Seasonal peak (week) | mem epi. threshold (week) | aedseo seasonal onset (week) | No. weeks between seasonal onset vs. mem epi. threshold | No. weeks with significant growth | No. weeks with no significant growth |
|---|---|---|---|---|---|---|---|
| J | ILI | 8 | 48 | 48 | 0 | 0 | 0 |
| O | RSV | 1 | 47 | 47 | 0 | 0 | 0 |
| P | RSV | 5 | 5 | 5 | 0 | 0 | 0 |

Table S3-3. Shows three data sources with the time (weeks) where aedseo estimates seasonal onset and mem crosses its epidemic threshold at the same week.

#### Supplement S4

It is important to be able to detect significant growth in the weekly observations to be able to alert that the circulation of the pathogen has started. A sensitivity analysis was conducted for influenza and RSV for the 21 European countries to select the optimal window parameter for the model. It was not done for ARI and ILI, as these are composite measures that are not expected to show a seasonal pattern dominated by a single wave. The *estimate_disease_threshold* function in the aedseo package was applied on each data source with default arguments and varying window sizes (3-8). This was done to investigate how the estimated disease-specific threshold is influenced by the window size.

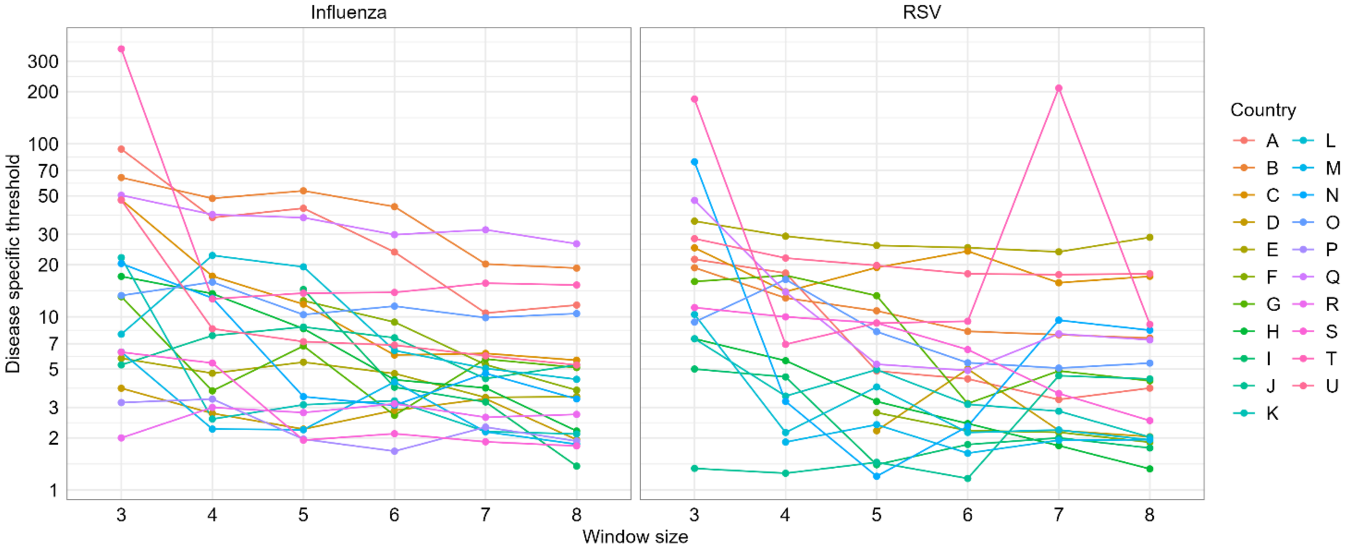


Figure S4. Shows the disease-specific threshold estimated by the *estimate_disease_threshold* function in the aedseo package as a function of window size for influenza and RSV sources from the 21 selected countries.

As aedseo uses five as the default window size, five is selected as the reference window size for comparison of the other window sizes (Fig. S4). When increasing the window size from the reference, the disease-specific threshold decreases and contrary it increases if decreasing the window size (Table S4-1). If the disease-specific threshold is too high the seasonal onset might be called too late in the season, whereas if it is too low we might have an early false alarm. Accordingly, the next natural step was to investigate how the combination of window size and disease-specific threshold actually influence the seasonal onset.

| Window | Disease | Threshold value shift median and IQR | Seasonal onset shift median and IQR |
|---|---|---|---|
| 3 | Influenza | 6.3 (1.43 - 17.88) | 0.5 (0 - 2) |
| 4 | Influenza | 0.52 (-0.84 - 3.33) | 0 (0 - 1) |
| 5 | Influenza | Reference | Reference |
| 6 | Influenza | -0.76 (-5.82 - 0.17) | 0 (-1 - 1) |
| 7 | Influenza | -1.23 (-5.69 - (-0.06)) | 0 (-3 - 0.5) |
| 8 | Influenza | -1.92 (-8.62 - (-0.14)) | 0 (-3.5 - 0.25) |
| 3 | RSV | 6.09 (2.69 - 11.57) | 0.5 (0 - 3.25) |
| 4 | RSV | 2.03 (-0.49 - 3.4) | 0 (-0.75 - 0) |
| 5 | RSV | Reference | Reference |
| 6 | RSV | -0.73 (-1.95 (-0.02)) | 0 (-1 - 0) |
| 7 | RSV | -1.54 (-2.62 - 0.31) | 0 (-0.25 - 1.75) |
| 8 | RSV | -1 (-2.47 - 0.1) | 0 (-1 - 2) |

*Table S4-1. Includes information about the median and IQR shift in the disease-specific threshold and seasonal onset when increasing/decreasing the window size from baseline (window size = 5), grouped by influenza and RSV from the 21 European countries.*

The *seasonal_onset* function from the aedseo package was used with default parameters and varying the window size with its corresponding disease-specific threshold (Fig. S4). Table S4-1 shows that the seasonal onset is in general called slightly later when using a smaller window size than baseline, and in contrary it is called slightly earlier or at the same time when using a bigger window size, which coincides with the disease-specific threshold being very low when increasing window size (Fig. S4-1). It is worth noting that the median shift in seasonal onset is zero weeks for most window sizes. Thus, the method is not that sensitive to small changes in window size.

| Window | Disease | Number of weeks between onset from aedseo to mem median and IQR | Number of significant weeks between onset from aedseo to mem median and IQR | Number of non-significant weeks between onset from aedseo to mem median and IQR |
|---|---|---|---|---|
| 3 | Influenza | 2 (1 - 6) | 2 (1.75 - 5) | 0 (0 - 3) |
| 4 | Influenza | 5 (3 - 10) | 3 (3 - 6) | 0 (0 - 3) |
| 5 | Influenza | 6 (2 - 11) | 4 (2 - 6.5) | 0 (0 - 3.5) |
| 6 | Influenza | 7 (3.5 - 11) | 5 (3.25 - 6.75) | 2.5 (0 - 3.75) |
| 7 | Influenza | 7 (4 - 12.5) | 5.5 (3.25 - 7) | 3 (0 - 7) |
| 8 | Influenza | 8 (6 - 12.5) | 5 (2.5 - 7.5) | 3 (1 - 7) |
| 3 | RSV | 2 (0 - 4) | 3 (2 - 3.75) | 0 (0 - 0) |
| 4 | RSV | 4 (2.75 - 7.5) | 4 (3 - 7.25) | 0 (0 - 0) |
| 5 | RSV | 4 (2 - 10) | 4 (3 - 7.25) | 0 (0 - 1.5) |
| 6 | RSV | 4 (3 - 10) | 4.5 (3.75 - 7.25) | 0 (0 - 2.25) |
| 7 | RSV | 4 (2 - 6) | 4 (3 - 6) | 0 (0 - 0) |
| 8 | RSV | 4 (2 - 7) | 4 (3.5 - 7) | 0 (0 - 0) |

Table S4-2. Includes information (median and IQR) of the number of weeks between seasonal onset detection by aedseo and reaching the epidemic threshold by mem. Comparations are made for window sizes 3-8 and estimations from the 21 European countries are grouped by influenza and RSV.

A sensitivity analysis was conducted for influenza and RSV across the 21 countries, by counting the number of weeks between when aedseo detects seasonal onset to when mem reaches the epidemic threshold and vice versa, while varying the aedseo window size between 3-8 weeks (Table S4-2). Across all window sizes, the median onset detected by aedseo occurred earlier than detected by mem. For influenza, results show that increasing the window size leads to a larger time gap between the two methods. For RSV, the time gap remains almost unchanged across window sizes. For both influenza and RSV, the three-week window gap is markedly small, which likely increases the risk of false detection.

For influenza, the number of significant weeks increase steadily from window size 3-5, without notable changes in the non-significant weeks. For window sizes 6-8 the number of non-significant weeks start to increase, implying that overly long windows begin to dilute early growth signals. For RSV, there are no notable changes in the number of significant and non-significant weeks across all window sizes. Consequently, selecting a window that is neither too short nor too long provides a better balance between early responsiveness and robustness to random variation. This trade-off appears more pronounced for influenza, while RSV shows little sensitivity to the chosen window size.

Based on literature [11] very short windows detect significant growth earlier but are dominated by sampling variability, often masking the underlying temporal pattern. Very long windows suppress that noise, yet they might postpone detection until the season is already well under way. This bias-variance trade-off is well documented [11], and our own sensitivity analysis shows the same for influenza, whereas RSV remained largely unaffected by window size (Table S4-2). A simulation study using the *generate_seasonal_data* function from the aedseo package (data not shown) confirmed these findings, with 200 simulations of 15 consecutive seasons yielding consistent results. Considering these findings and given that observations are reported weekly, we adopted a five-week window, which is long enough to smooth random fluctuations and reveal sustained trends, while remaining sensitive to early emerging changes.

#### Supplement S5

When estimating the high breakpoint (between the high and very high burden levels) the goal is for it to only be exceeded in truly extreme seasons. To reflect this best, a sensitivity analysis was conducted for influenza and RSV for the 21 European countries to select the optimal number of peak observations from each season by running the aedseo analysis with varying peak observations (1-8) per season.

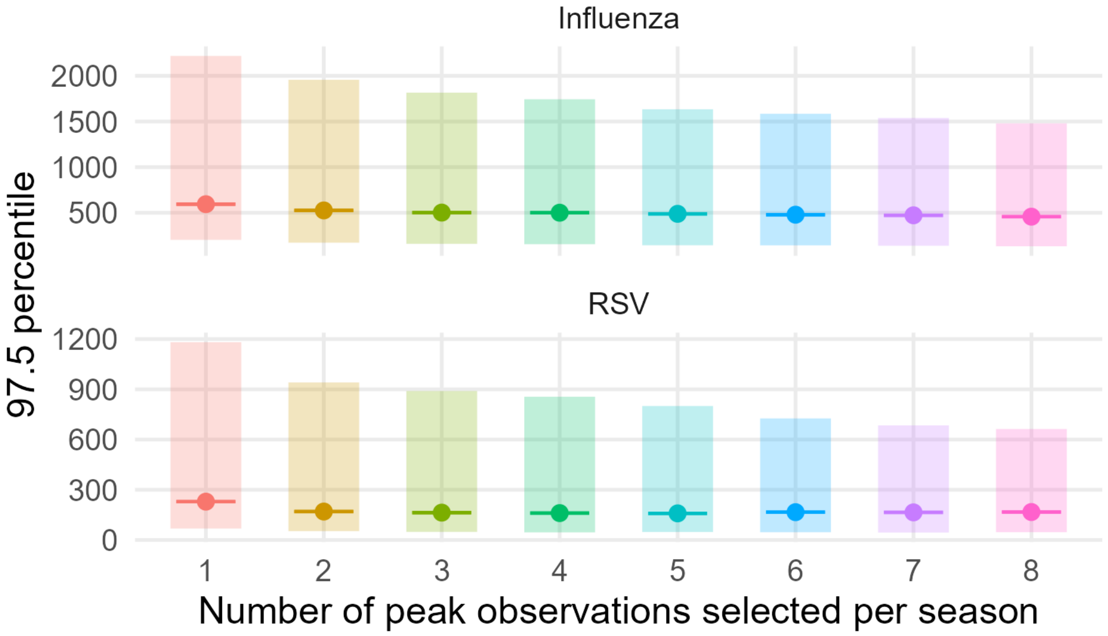


Figure S5-1. Distribution of estimated 97.5th percentiles (median and IQR across all seasons for influenza and RSV in 21 European countries) versus the number of peak observations used in the high breakpoint estimation by aedseo.

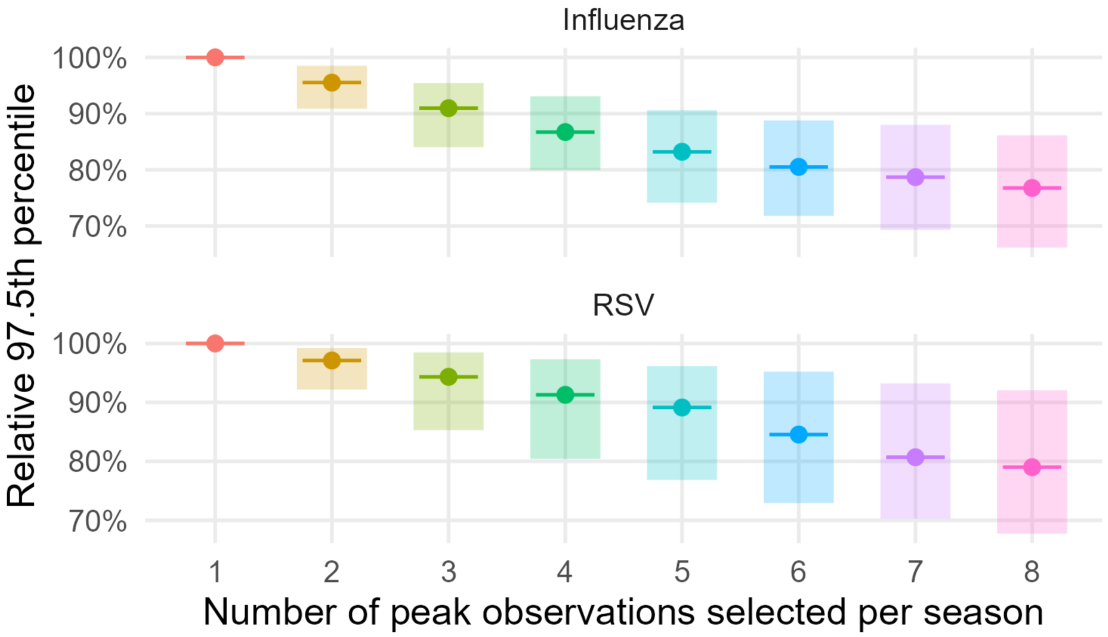


Figure S5-2 Relative 97.5th percentile (median and IQR across all seasons for influenza and RSV in 21 European countries) to the 97.5 percentile when using one peak observation in the high breakpoint estimation by aedseo versus the number of peak observations selected per season (2-8).

When varying the number of peak observations used to estimate the 97.5th percentile using aedseo, we observe a consistent pattern across both influenza and RSV (Figures S5-1 and S5-2), where increment in number of peak observations leads to lower percentiles, hence a lower high breakpoint. From figure S5-2 it is evident that the 97.5th percentile decreases steadily when increasing the number of peak observations included in the estimation. Including more peaks therefore lowers the estimated 97.5th percentile (risking underestimation of true high-intensity seasons), while too few peaks lead to bias from e.g. extreme years or more testing. When including two or three peak observations the median 97.5th percentile is still above 90% of the 97.5th percentile estimation when using one peak observation for both influenza and RSV.

Despite substantial heterogeneity in detection definitions, reporting systems, and data quality across seasons and diseases across 21 European countries, the same systematic pattern was observed. This trend was confirmed using the *generate_seasonal_data* function from the aedseo package (data not shown), with 200 simulations of 15 consecutive seasons. This indicates that the underlying behaviour of the method is robust even under real-world surveillance variability. To balance this bias-variance trade-off three peak observations from each season are used in the aedseo method.
